# Sex shapes plasma molecular signatures of dementia risk across *APOE* ε4 strata

**DOI:** 10.64898/2026.11.02.26366836

**Authors:** Sara Calhas, Yue Liu, Sheena Waters, Dorsa Abdolkarimi, Cristina Legido-Quigley, Oleg Blyuss, Charles R Marshall, Petroula Proitsi

## Abstract

Sex and *APOE* ε4 influence dementia risk, but whether its molecular predictors differ across sex and *APOE* ε4 strata remains unclear. We investigated plasma proteomic and metabolomic signatures predicting future dementia in 43,759 UK Biobank participants across four sex-by-*APOE* ε4 groups. The association between *APOE* ε4 carrier status and dementia differed by sex (interaction p = 0.027). Machine-learning models achieved higher discrimination in *APOE* ε4 carriers (AUC 0.85–0.90) than non-carriers (AUC 0.74–0.76), with the highest performance in female carriers. Most top molecular predictors were subgroup-specific, including C7, CDON and EDIL3 in female carriers and KLK4, MMP12 and TGFBI in male carriers. When UK Biobank models were applied to the Global Neurodegeneration Proteomics Consortium, 12/17 available top predictors in female carriers and 10/16 in male carriers remained highly ranked. These findings identify distinct molecular signatures of dementia risk across sex and *APOE* ε4 strata, supporting stratified biomarker discovery.

## Main

Dementia is a leading cause of disability and mortality worldwide, affecting over 50 million people, with prevalence projected to exceed 150 million by 2050 (1).

Beyond age, sex and *APOE* ε4 genotype are two of the strongest risk factors for late-onset Alzheimer’s disease (AD) (2). *APOE* ε4 carriers have increased dementia risk, with homozygotes exhibiting up to 15-fold higher AD risk than APOE ε3 homozygotes (3). An APOE-stratified genome-wide association study further identified genetic associations that differ according to *APOE* carrier status, suggesting distinct genetic mechanisms contributing to AD (4). Females are disproportionately affected by dementia (2,5), with some studies reporting greater *APOE* ε4-associated risk in females than males (6). Emerging evidence further suggests that sex may modify disease progression, biomarker trajectories (7–10), molecular profiles associated with AD (11), and potentially therapeutic response in AD (12). Finally, *APOE* ε4 carrier status, particularly homozygosity, is associated with increased risk of amyloid-related imaging abnormalities during anti-amyloid treatment (13).

Nevertheless, dementia risk prediction studies have largely focused on population-level models, with less attention to the molecular heterogeneity underlying risk across sex and *APOE* ε4 subgroups. Identifying subgroup-specific molecular signatures before clinical dementia onset may clarify heterogeneity in neurodegeneration, provide insights into the biological basis of variation in dementia risk, and inform future precision prevention strategies (14).

High-throughput plasma proteomics and metabolomics enable minimally invasive investigation of molecular changes associated with neurodegeneration. Machine learning can further model complex relationships within high-dimensional biological data and is increasingly applied to dementia research (15–18). A pioneering large-scale study demonstrated the potential of plasma proteomics and machine learning for predicting incident dementia (19). However, whether the molecular features driving dementia prediction differ across sex and *APOE* ε4 strata remains insufficiently characterised. Furthermore, external validation, cross-platform transportability, and biological interpretation of machine learning-derived molecular signatures remain limited.

Using UK Biobank (UKB) (20) as a prospective discovery cohort, we integrated plasma proteomics and metabolomics within sex and *APOE* ε4 strata to identify molecular signatures of future dementia risk. We evaluated the cross-cohort and cross-platform transportability of the identified proteomic signatures in the Global Neurodegeneration Proteomics Consortium (GNPC) (21), a clinically enriched cohort using a distinct proteomic platform. SHAP-based model interpretation, pathway enrichment, and association analyses were used to characterise the identified molecular signatures and explore their biological context.

## Results

### Cohort Characteristics (UK Biobank)

Participants with dementia were significantly older at baseline (median 66 years) compared with controls (median 58 years, p < 0.001). Years of education were lower among dementia cases (median 11 vs. 12 years, p < 0.001). The sex distribution differed modestly between groups, with males representing a greater proportion of dementia cases than controls (49.3% vs. 45.7%, p = 0.018). *APOE* ε4 genotype distribution differed substantially between groups, with *APOE* ε4 homozygotes accounting for 18.6% of incident dementia cases compared with 2.5% of controls (p < 0.001). Ethnic composition also differed between participants with and without dementia (p < 0.001). BMI category distributions did not differ between groups (p = 0.130; **Supplementary Table 1**).

When stratified by sex and *APOE* ε4 carrier status (**Table 1**), the largest subgroup was female non-carriers (n = 16,476 controls, 221 cases), followed by male non-carriers (n = 14,049 controls, 256 cases), female carriers (n = 6,643 controls, 364 cases), and male carriers (n = 5,437 controls, 313 cases). Among females, 5.2% of *APOE* ε4 carriers and 1.3% of non-carriers had dementia. Among males, 5.4% of carriers and 1.8% of non-carriers had dementia.

**Table 1.** Counts and proportions of dementia cases and controls stratified by sex and *APOE* ε4 carrier status.

| **Sex** | ***APOE* ε4 status** | **Dementia cases (n, %)** | **Controls (n, %)** | **Total (n)** |
|---|---|---|---|---|
| Female | Non-carrier | 221 (1.3%) | 16,476 (98.7%) | 16,697 |
| Female | Carrier | 364 (5.2%) | 6,643 (94.8%) | 7,007 |
| Male | Non-carrier | 256 (1.8%) | 14,049 (98.2%) | 14,305 |
| Male | Carrier | 313 (5.4%) | 5,437 (94.6%) | 5,750 |

### Baseline epidemiological associations (UK Biobank)

In multivariable logistic regression analyses (**Figure 1**) adjusted for age, years of education, BMI category, and ethnicity, the magnitude of the association between *APOE* ε4 carrier status and dementia differed significantly by sex (*APOE* ε4 carrier × sex interaction OR = 0.76, 95% CI 0.60–0.97, p = 0.027), with a stronger association observed in females than males. *APOE* ε4 carrier status was strongly associated with increased dementia risk in both females (OR = 4.11, 95% CI 3.46–4.89, p < 0.001) and males (OR = 3.12, 95% CI 2.63–3.71, p < 0.001).

**Figure 1.**
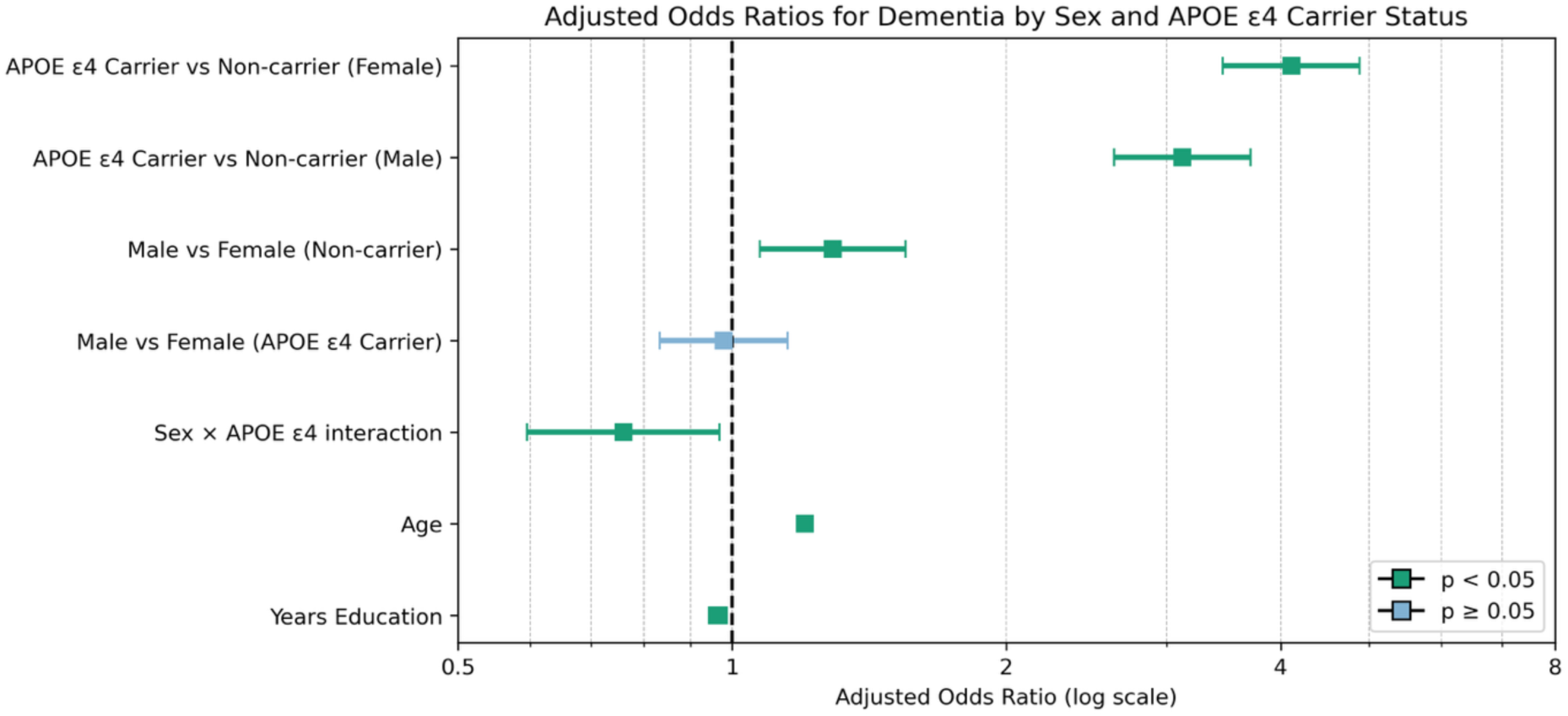
Forest plot of adjusted odds ratios from multivariable logistic regression, showing associations of *APOE* ε4 status, sex, their interaction, and covariates with dementia risk.

Among *APOE* ε4 non-carriers, males had higher dementia risk than females (male vs female OR = 1.29, 95% CI 1.07–1.55, p = 0.007), whereas no significant sex difference was observed among *APOE* ε4 carriers (male vs female OR = 0.98, 95% CI 0.83–1.15, p = 0.795). Full results are provided in **Supplementary Tables 7 and 8**. Multicollinearity diagnostics indicated low collinearity between predictors, with all non-intercept VIF values below 3 (**Supplementary Table 9**).

### Feature Selection (Boruta)

Boruta feature selection identified between 22 and 71 stable features across the eight models, comprising four sex- and *APOE* ε4-stratified subgroups (female carriers, female non-carriers, male carriers, and male non-carriers), each evaluated with and without age as a predictor (≥40% stability threshold; **Supplementary Tables 10 to 17**). The highest numbers of stable features were identified in male and female *APOE* ε4 carriers (65-71, depending on age inclusion), whereas the fewest were observed in female and male non-carriers (22 in models including age). Five proteins reached the predefined stability threshold across all groups: EDA2R, ELN, GDF15, GFAP, and NEFL **(Supplementary Table 18)**.

### Machine learning model performance

Model performance varied across sex and *APOE* ε4 strata (**Supplementary Table 19, Figure 2**). Female carriers achieved the highest discrimination (AUC 0.88–0.90), with recall of 0.72-0.78 and specificity of 0.85-0.87. Male carriers also showed strong discrimination (AUC 0.85 in both models), with specificity of 0.85-0.86 and recall of 0.69-0.71. Performance was lower in female (AUC 0.75–0.76) and male non-carriers (AUC 0.74–0.76).

**Figure 2.**
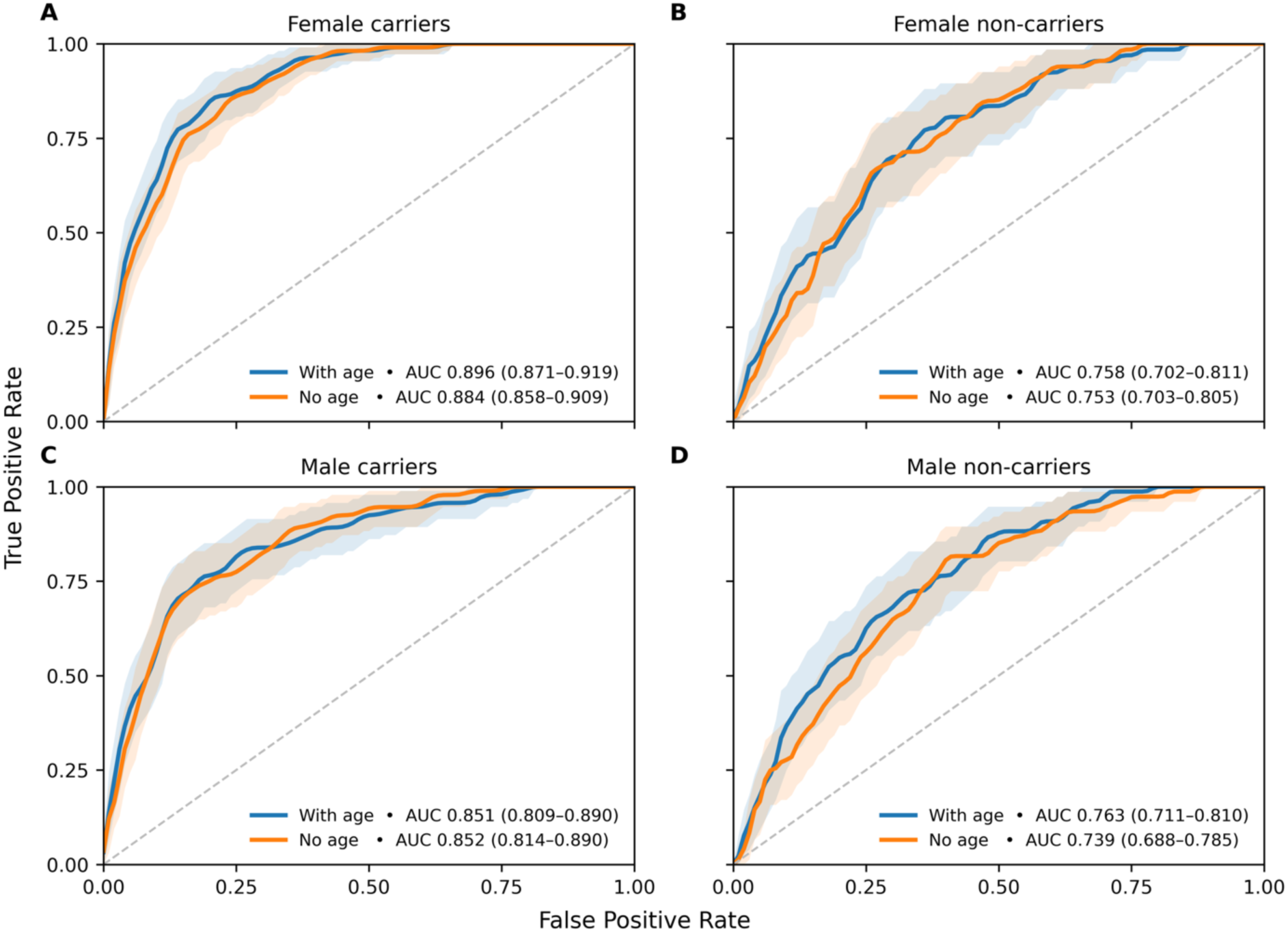
Receiver operating characteristic (ROC) curves with 95% confidence intervals for XGBoost dementia classifiers stratified by sex and *APOE* ε4 status. Panels show female *APOE* ε4 carriers (A), female non-carriers (B), male *APOE* ε4 carriers (C), and male non-carriers (D). Models including age (blue) incorporated chronological age alongside proteomic and metabolomic predictors, whereas models excluding age (orange) used molecular predictors only. Inclusion of age resulted in modestly higher AUCs in some subgroups, although discrimination remained broadly preserved in models excluding age. Female *APOE* ε4 carriers achieved the highest overall discrimination, whereas non-carrier groups demonstrated lower predictive performance.

Inclusion of age produced modest increases in AUC across subgroups (**Figure 2**, **Supplementary Table 19**). Paired stratified bootstrap comparisons showed small increases in AUC for female carriers (ΔAUC = 0.013, 95% CI 0.003–0.022) and male non-carriers (ΔAUC = 0.024, 95% CI 0.006–0.040), whereas confidence intervals included zero for female non-carriers (ΔAUC = 0.004, 95% CI −0.019–0.028) and male carriers (ΔAUC = −0.001, 95% CI −0.013–0.010; **Extended Data Figure 2**, **Supplementary Table 20**). Overall, differences in discrimination between models with and without age were small.

Following sigmoid calibration, Brier scores ranged from 0.013 to 0.046, with improved agreement between predicted and observed risks across subgroups (**Extended Data Figure 3, Supplementary Table 21).**

A targeted sensitivity analysis was conducted in a subset of 2,000 female *APOE* ε4 carriers to assess the robustness of model discrimination to potential data leakage during preprocessing. Using models without age as a predictor, we compared the original preprocessing approach with a revised strategy in which KNN imputation was fitted exclusively on the training data. Model discrimination remained comparable (AUC 0.883 versus 0.893, respectively), with highly correlated predictions (r = 0.979; **Supplementary Table 21a**).

### Feature Importance (SHAP)

To identify the features driving model predictions, we examined SHAP values for each subgroup-specific model. The relative ranking of predictors was highly consistent between models with and without age (**Supplementary Table 27**). Given the broadly preserved predictive performance and consistency in feature rankings between models with and without age, SHAP results from no-age models were used for primary molecular interpretation. This allowed us to examine the contributions of molecular predictors without chronological age entering the models directly, although age-related biological variation may still be reflected in the selected proteins.

Analysis of the top 20 SHAP-ranked features revealed both shared and subgroup-specific predictors, indicating substantial heterogeneity in predictive molecular signatures across sex and *APOE* ε4 strata (**Extended Data Figure 4**; **Supplementary Tables 22 –26**).

Among the top 20 predictors, 11 were specific to female carriers and 10 to male carriers, compared with 14 in each non-carrier group (**Figure 3**, **Supplementary Table 26**). Despite this greater feature diversity among non-carriers, predictive performance was consistently higher in *APOE* ε4 carriers.

**Figure 3.**
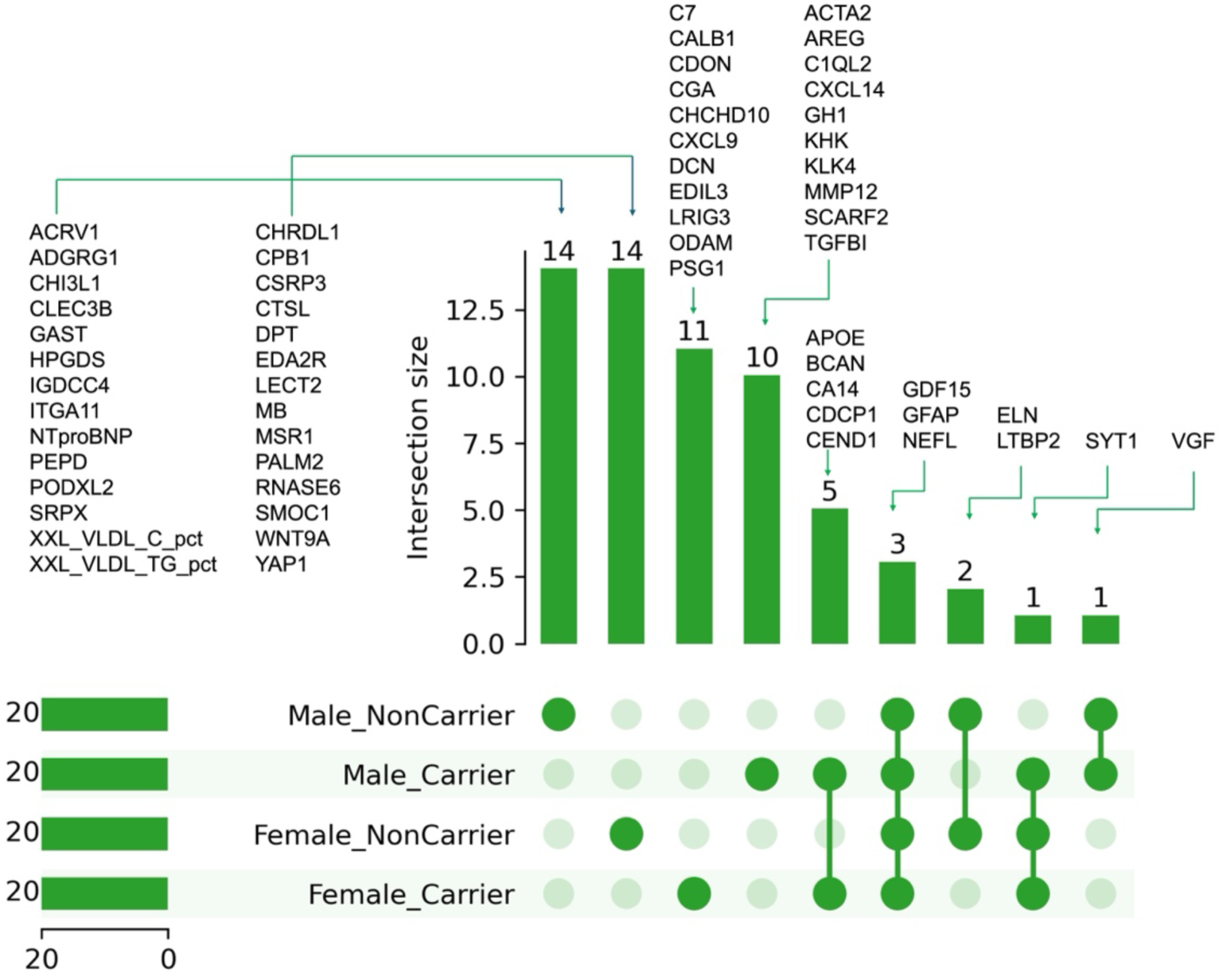
Overlap of top 20 SHAP-ranked predictors across sex– *APOE* ε4 strata from the no-age XGBoost models. UpSet plot showing shared and subgroup-specific predictors among female *APOE* ε4 carriers, male *APOE* ε4 carriers, female non-carriers, and male non-carriers. Vertical bars represent the number of overlapping predictors within each subgroup combination, while connected dots indicate subgroup membership for each intersection. GFAP, NEFL and GDF15 were the only proteins shared across all four groups, whereas most predictors were subgroup-specific.

Overlap of top predictors across strata was limited (**Figure 3**). *APOE* ε4 carriers shared five proteins among their top-ranked features (BCAN, APOE, CA14, CEND1 and CDCP1), whereas non-carriers shared only LTBP2 and ELN. Additional overlaps included VGF in both male strata and SYT1 across female carriers, female non-carriers and male carriers. Across all four groups, GFAP, NEFL and GDF15 were the only proteins consistently represented among the top predictors.

Proteins dominated the top-ranked predictors, with two metabolomic measures (XXL_VLDL_C_pct and XXL_VLDL_TG_pct, representing the cholesterol and triglyceride content of extremely large VLDL particles) appearing among the top predictors only in male *APOE* ε4 non-carriers.

These findings suggest that the molecular features contributing to dementia prediction differ substantially across sex and *APOE* ε4 strata. Complete ranked SHAP feature importance lists for each subgroup are provided in **Supplementary Tables 22 –26**.

### SHAP-Based Gene Set Enrichment results

Contrast-based Reactome gene set enrichment analysis was used to investigate biological differences between female and male molecular signatures within *APOE* ε4 carrier and non-carrier strata (**Figure 4**).

**Figure 4.**
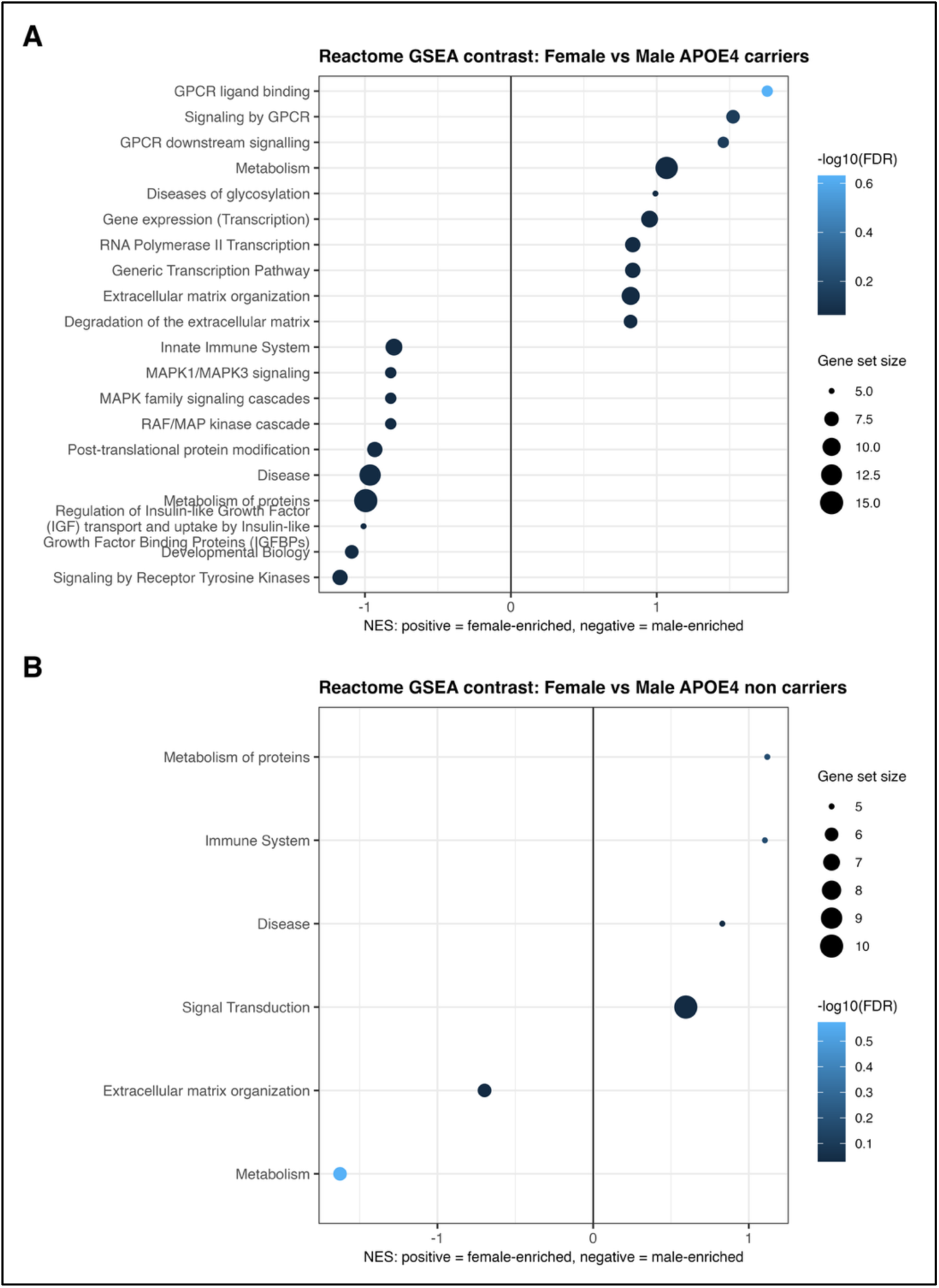
Reactome pathway enrichment analysis of sex-specific molecular signatures. Contrast-based Reactome GSEA comparing females and males within (A) *APOE* ε4 carriers and (B) *APOE* ε4 non-carriers. Positive normalised enrichment scores (NES) indicate female-enriched pathways, and negative NES values indicate male-enriched pathways. Point size reflects gene set size and colour represents enrichment significance (-log10 FDR-adjusted p-value).

Among *APOE* ε4 carriers, proteins contributing more strongly to prediction in females were enriched for GPCR ligand binding and downstream signalling, metabolism, cytokine signalling, gene transcription and extracellular matrix organisation. In contrast, proteins contributing more strongly to prediction in males were enriched for innate immune system processes, MAPK signalling cascades, receptor tyrosine kinase signalling, developmental biology, regulation of insulin-like growth factor transport and protein metabolism **( Figure 4A**).

A different pattern was observed among *APOE* ε4 non-carriers. Female-associated predictors were enriched for pathways related to signal transduction, immune system processes and protein metabolism. In contrast, male-associated predictors were enriched for extracellular matrix organisation and broader metabolic pathways (**Figure 4B**).

Although these findings did not survive correction for multiple testing and should therefore be considered exploratory, they suggest potentially distinct biological processes underlying the sex and *APOE* ε4-specific molecular signatures. Full enrichment results, including adjusted p-values, are provided in **Supplementary Tables 28 –31**.

### External validation cohort, proteomic coverage and classifier performance

Because the UKB models for *APOE* ε4 carriers demonstrated the strongest predictive performance (AUC 0.85–0.90), we evaluated whether their proteomic signatures replicated in GNPC. Metabolomic data were unavailable in GNPC. However, all predictors retained through Boruta feature selection in the UKB *APOE* ε4 carrier models were proteins, allowing the original models to be evaluated using proteomic measurements alone.

Compared with UKB, GNPC participants were older and enriched for AD cases (**Supplementary Table 32).** Of the model predictors identified in UKB, 55/68 in female carriers and 57/71 in male carriers were available in GNPC (**Supplementary Tables 33** and **34**).

The external validation framework provided a stringent test of molecular transportability given differences in cohort composition (population-based versus clinically enriched), disease phenotype and diagnostic context (incident all-cause dementia in UKB versus prevalent clinically diagnosed Alzheimer’s disease in GNPC), proteomic platform (Olink versus SomaScan), and cohort age structure.

Classifier performance was lower in GNPC than in UKB (**Figure 5A, B**). Prediction retained modest discrimination (AUC 0.63 for female carriers and 0.65 for male carriers), indicating partial transfer of the predictive signal. Full classifier results are provided in **Supplementary Table 35**.

**Figure 5.**
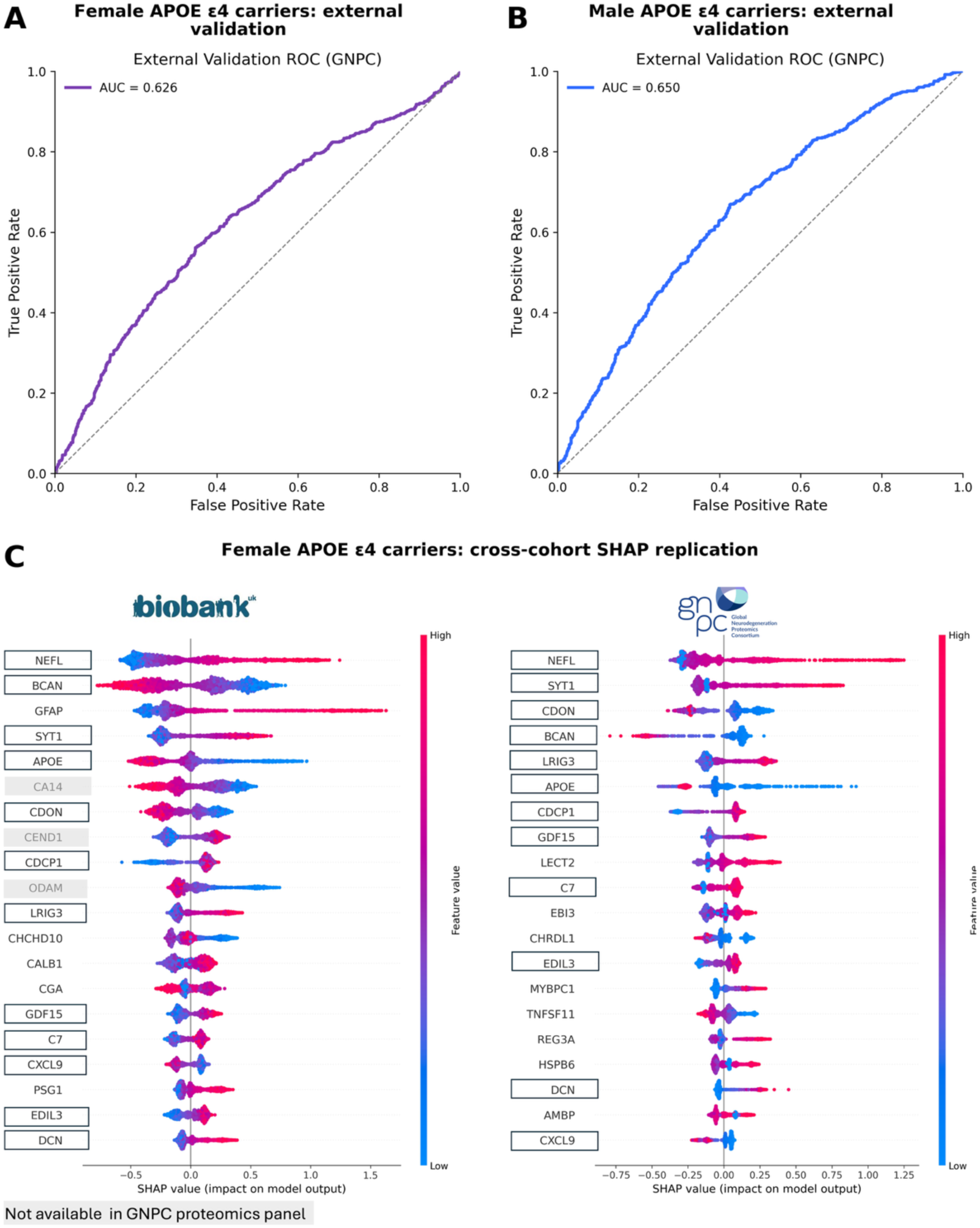
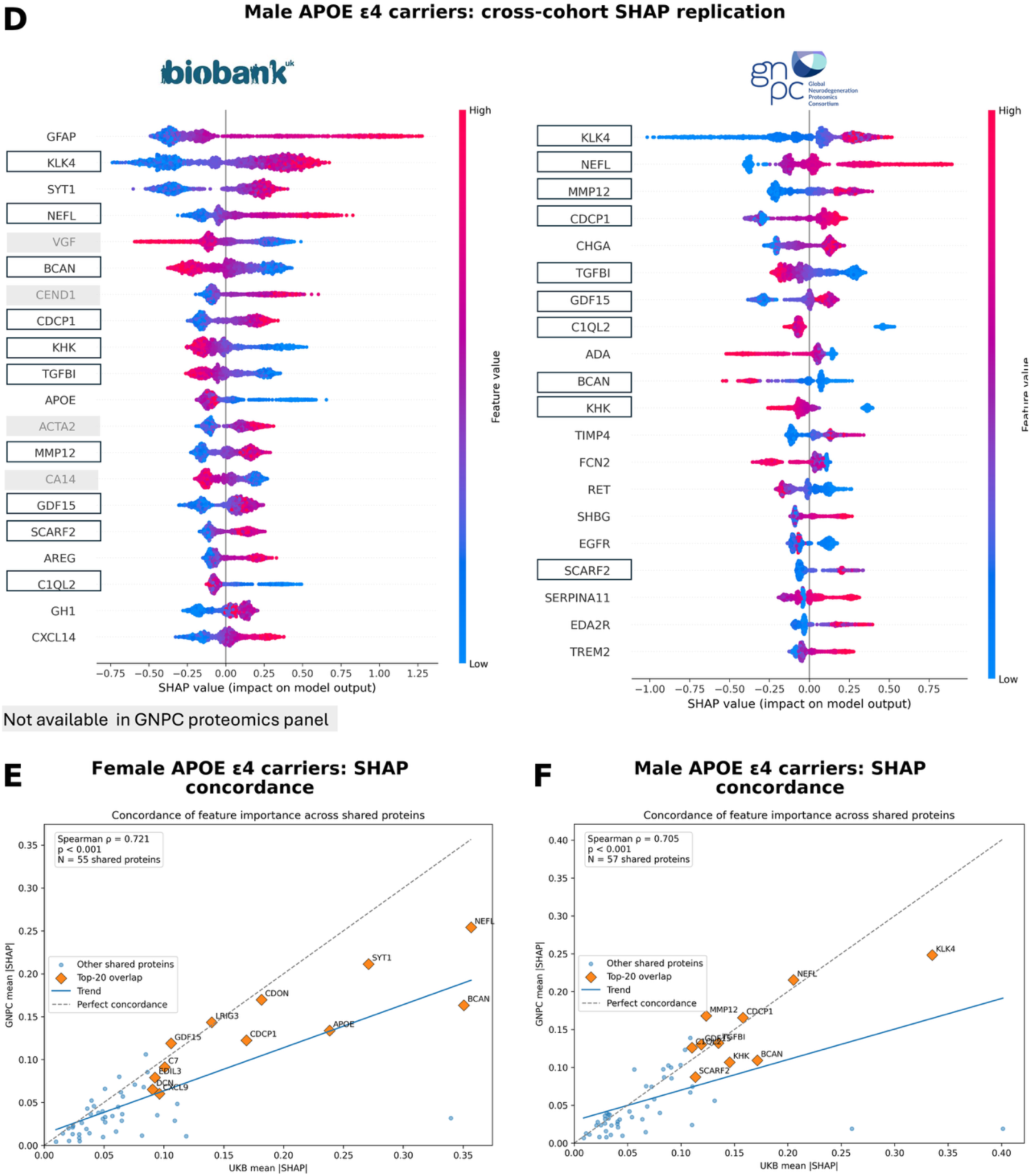
External validation and cross-cohort replication of *APOE* ε4 carrier models and proteomic signatures in GNPC. (A,B) Receiver operating characteristic (ROC) curves for external validation of UK Biobank-derived models in female (A) and male (B) *APOE* ε4 carriers. (C,D) Cross-cohort replication of the top 20 SHAP-ranked predictors in female (C) and male (D) *APOE* ε4 carriers. Proteins outlined in black were among the top 20 predictors in both cohorts; proteins shaded in grey were unavailable in GNPC. (E,F) Concordance of mean absolute SHAP values for shared proteins between UK Biobank and GNPC in female (E) and male (F) *APOE* ε4 carriers. Orange diamonds indicate proteins among the top 20 predictors in both cohorts and blue circles other shared proteins; dashed diagonal lines indicate perfect concordance.

### Cross-cohort replication of molecular predictors

Among female carriers, 12 of the 17 UKB top 20 predictors available in GNPC (70%) replicated among the GNPC top 20: NEFL, BCAN, SYT1, APOE, CDON, CDCP1, LRIG3, GDF15, C7, CXCL9, EDIL3 and DCN (**Figure 5C**).

Of the 11 predictors unique to female carriers in UKB (**Figure 3**), six replicated in GNPC (C7, CDON, CXCL9, DCN, EDIL3 and LRIG3). As ODAM was unavailable in GNPC, this represented 6/10 (60%) available female-specific predictors.

Among male carriers, 10 of the 16 UKB top 20 predictors available in GNPC (63%) replicated among the GNPC top 20: KLK4, NEFL, BCAN, CDCP1, KHK, TGFBI, MMP12, GDF15, SCARF2 and C1QL2 (**Figure 5D**).

Of the 10 predictors unique to male carriers in UKB, six replicated in GNPC (C1QL2, KHK, KLK4, MMP12, SCARF2 and TGFBI). As ACTA2 was unavailable, this represented 6/9 (67%) available male-specific predictors.

Full SHAP feature importance rankings and the classification of shared and sex-specific molecular predictors in GNPC are provided in **Supplementary Tables 36 –38**.

### Cross-cohort SHAP concordance

SHAP feature importance across shared proteins, calculated using the original UKB-trained models applied to GNPC, was strongly correlated between UKB and GNPC in both female (n = 55; Spearman ρ = 0.72, p < 0.001) and male (n = 57; ρ = 0.70, p < 0.001) *APOE* ε4 carriers (**Figure 5E, F**).

Several overlapping top-ranked proteins clustered close to the perfect concordance line in both female and male *APOE* ε4 carriers, indicating preservation of relative feature importance across cohorts. Among female *APOE* ε4 carriers, CDON, LRIG3, GDF15, C7 and EDIL3 demonstrated highly preserved SHAP importance. Similarly, among male *APOE* ε4 carriers, NEFL, CDCP1, TGFBI, GDF15, MMP12 and C1QL2 clustered near the concordance line.

### Associations of replicated proteins with dementia outcomes in *APOE* ε4 carriers (UKB)

Following cross-cohort evaluation in GNPC, the 12 proteins replicated among female carriers and 10 among male carriers were taken forward for association analyses with all-cause dementia, Alzheimer’s disease (AD), and vascular dementia (VD). With the additional inclusion of GFAP in both groups and APOE and SYT1 in males, 13 proteins were evaluated in each sex. Analytic sample sizes varied by outcome, ranging from 6,700–7,007 female and 5,511–5,750 male *APOE* ε4 carriers, with outcome-specific case and control counts provided in **Supplementary Table 39**.

### Female *APOE* ε4 carriers

Of the top 13 proteins tested, 11 were significantly associated with at least one dementia outcome after FDR correction (**Figure 6A**; **Supplementary Table 40**). Specifically, 11 proteins were significantly associated with all-cause dementia, 7 with AD, and 6 with vascular dementia. Four proteins (GFAP, NEFL, SYT1 and GDF15) were consistently associated across all three outcomes, indicating substantial overlap in biomarker profiles across dementia phenotypes.

**Figure 6.**
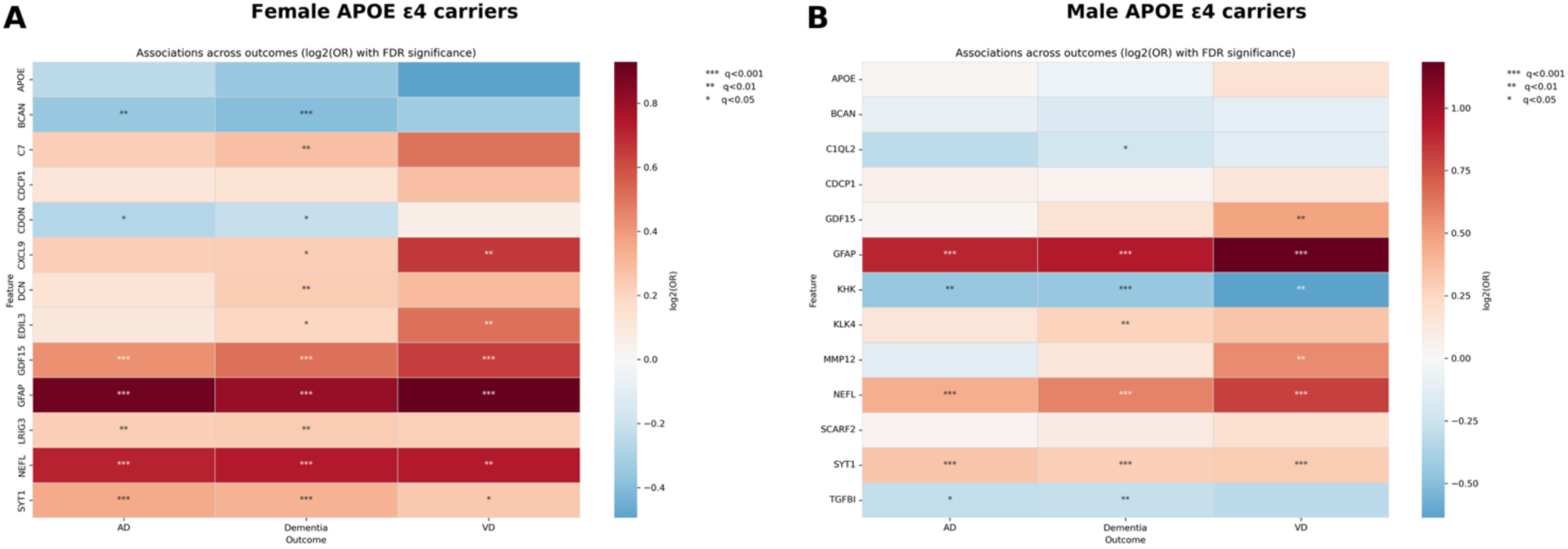
Sex-stratified associations of replicated *APOE* ε4 carrier proteins with dementia-related outcomes in UK Biobank. Heatmaps show log2 odds ratios from multivariable logistic regression analyses in female (A) and male (B) *APOE* ε4 carriers for Alzheimer’s disease (AD), all-cause dementia and vascular dementia (VD). Models were adjusted for age, education, *APOE* ε4 allele count, BMI category and ethnicity. Positive values indicate increased odds of dementia associated with higher protein levels, whereas negative values indicate inverse associations. Asterisks indicate statistical significance after false discovery rate correction (\**q* < 0.05, \*\**q* < 0.01, \*\*\**q* < 0.001). Colour scales are shown separately for each sex.

GFAP and NEFL demonstrated the strongest positive associations across outcomes (**Supplementary Table 40, Extended Data Figures 5** and 6). For example, higher GFAP levels were associated with increased odds of all-cause dementia (OR=1.74, 95% CI 1.53-1.98, q<0.001), of AD (OR=1.87, 95% CI 1.60–2.19, q<0.001), and of VD (OR=1.90, 95% CI 1.49-2.43, q<0.001). Higher SYT1 and GDF15 levels were also consistently associated with increased odds of all dementia-related outcomes.

In contrast, lower BCAN and CDON levels were associated with increased odds of AD and all-cause dementia. These inverse associations were consistent with SHAP patterns in UKB and GNPC, where lower levels contributed towards higher dementia risk predictions.

Patterns of statistical significance varied across dementia outcomes. BCAN, CDON, and LRIG3 showed significant associations with AD and all-cause dementia, but not vascular dementia, whereas CXCL9 and EDIL3 showed significant associations with all-cause dementia and vascular dementia. C7 and DCN reached statistical significance only for all-cause dementia.

### Male *APOE* ε4 carriers

Of the 13 proteins tested, 9 (C1QL2, GDF15, GFAP, KHK, KLK4, MMP12, NEFL, SYT1 and TGFBI) were significantly associated with at least one dementia-related outcome after FDR correction (**Figure 6B; Supplementary Table 40**).

Four proteins (GFAP, NEFL, SYT1 and KHK) were associated with all three dementia outcomes. As observed in female carriers, GFAP and NEFL had the strongest positive associations, with higher levels associated with increased odds of dementia-related outcomes (**Extended Data Figures 7 and 8**).

Patterns of statistical significance also varied across outcomes in male carriers. TGFBI showed significant associations with all-cause dementia and AD. C1QL2 and KLK4 reached statistical significance only for all-cause dementia, while GDF15 and MMP12 reached significance only for vascular dementia.

The directionality of the logistic regression associations was consistent with SHAP analyses from both the UK Biobank and GNPC machine-learning models. Higher circulating levels of GFAP, NEFL, GDF15, KLK4, SYT1 and MMP12 were associated with increased odds of dementia-related outcomes, whereas lower C1QL2, KHK and TGFBI levels were associated with increased dementia risk.

Full regression outputs, including nominally significant associations, are provided in Supplementary Table 40.

### Associations of shared proteins across all strata in *APOE* ε4 non-carriers (UKB)

To complement the analyses in *APOE* ε4 carriers, we examined associations of the shared molecular predictors GFAP, GDF15 and NEFL, together with SYT1, across all-cause dementia, AD and VD in female and male *APOE* ε4 non-carriers. Analytic sample sizes varied by outcome, ranging from 16,516–16,697 female and 14,117–14,305 male *APOE* ε4 non-carriers. GFAP and NEFL were significantly associated with all three dementia outcomes in both female and male non-carriers. GDF15 was significantly associated with all three outcomes in female non-carriers and with all-cause dementia and AD, but not VD, in male non-carriers. SYT1 was significantly associated with all-cause dementia and AD, but not VD, in female non-carriers, while no significant associations were observed in male non-carriers. Full sample counts and association results are provided in **Supplementary Tables 41and 42**.

## Discussion

In this study, we applied a stratified machine learning framework integrating plasma proteomics and metabolomics to characterise sex–*APOE* ε4-specific molecular signatures predictive of future dementia risk. To our knowledge, this is among the first studies combining sex-stratified *APOE* ε4 analyses with multimodal omics, interpretable machine learning, and external cross-platform validation to investigate molecular heterogeneity underlying dementia risk. **Figure 7** integrates the principal molecular risk patterns identified across analytical approaches.

**Figure 7.**
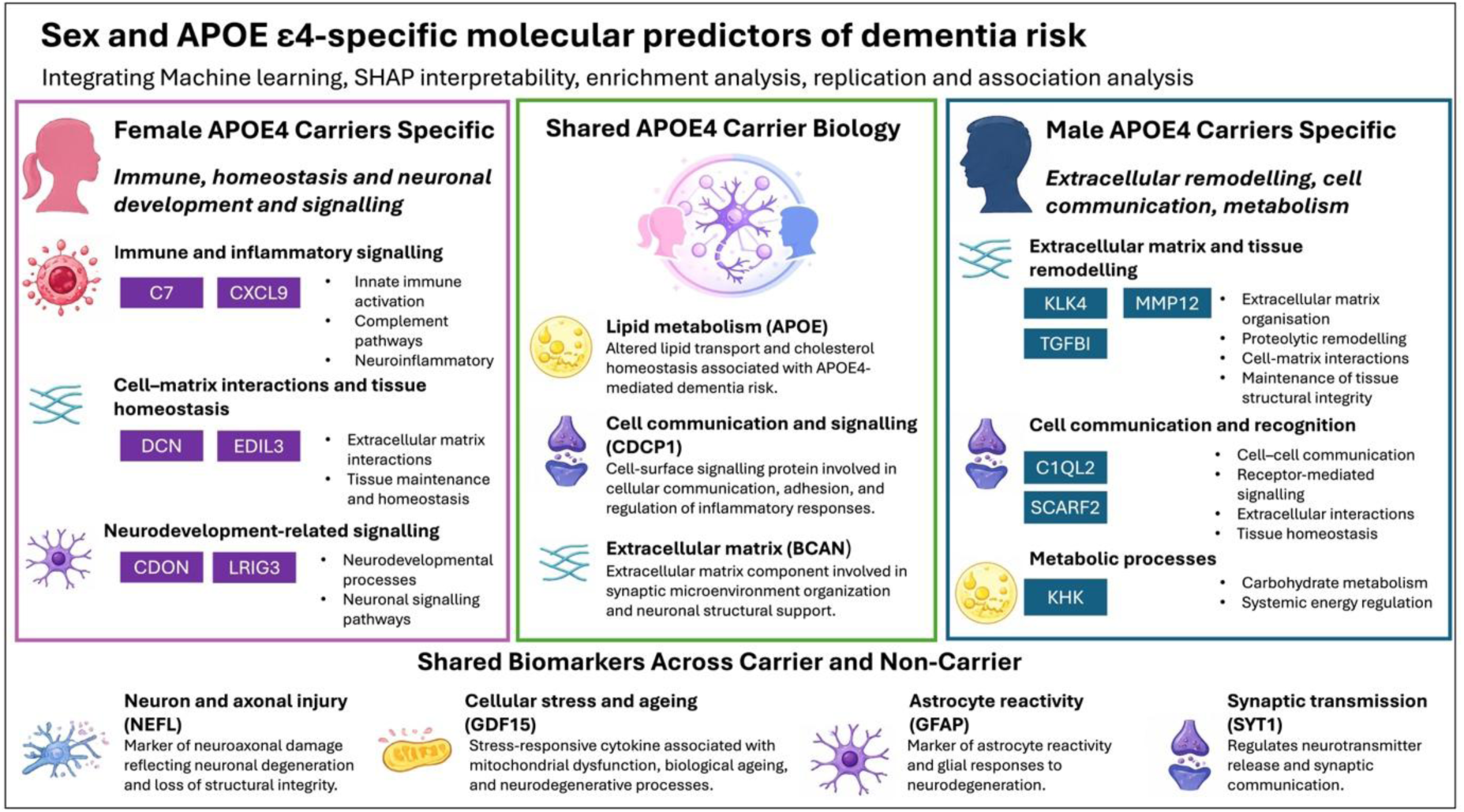
Integrated biological synthesis of sex- and *APOE* ε4-specific molecular signatures of dementia risk. Molecular features identified across stratified machine-learning, cross-cohort replication, and association analyses are summarised into female *APOE* ε4 carrier-specific pathways, shared *APOE* ε4-associated biology, male *APOE* ε4 carrier-specific pathways, and biomarkers shared across carrier and non-carrier populations.

Three principal findings emerged. First, molecular signatures differed substantially across sex– *APOE* ε4 strata, revealing both shared and subgroup-specific molecular predictors. Second, *APOE* ε4 carriers demonstrated stronger molecular predictive signals than non-carriers, with female carriers showing the highest model discrimination. Third, several proteomic predictors and SHAP-derived feature importance rankings were preserved in GNPC despite differences in cohort composition, disease stage, outcome definition and proteomic platform. These findings support partial cross-cohort transportability of molecular predictor importance.

One notable observation was the stronger predictive performance among *APOE* ε4 carriers compared with non-carriers, accompanied by a larger number of stable Boruta-selected features. This may reflect more consistent molecular associations with dementia risk among carriers, potentially driven by a greater contribution of Alzheimer’s disease pathology to dementia outcomes in this group. This interpretation is biologically plausible given the involvement of *APOE* ε4 in amyloid accumulation (22,23), neuroinflammation (24,25), lipid dysregulation (26), synaptic dysfunction (27), and blood–brain barrier impairment (28). However, differences in dementia case numbers, case composition and statistical power may also contribute to the observed differences in model performance and feature selection.

Distinct molecular signatures emerged among female and male *APOE* ε4 carriers, reflecting differences in immune signalling, tissue remodelling, neuronal communication and metabolic processes. These patterns were supported by the integration of machine learning feature importance, exploratory pathway enrichment, cross-cohort preservation of selected predictors and conventional association analyses.

Several female *APOE* ε4 carrier-specific predictors demonstrated substantial preservation across cohorts, including C7, EDIL3, CDON, LRIG3, CXCL9 and DCN. C7, a component of the complement system, implicates inflammatory processes relevant to AD pathology (29,30), while EDIL3 (31,32) and DCN (33) point towards vascular regulation and extracellular matrix organisation. CXCL9 (34), a chemokine involved in immune cell recruitment and inflammatory signalling, further supports the immune-related component of this signature. CDON (35) and LRIG3 (36), which participate in developmental and growth factor signalling, further point towards neurodevelopmental processes and cell-to-cell communication. Together, these findings highlight a female *APOE* ε4 carrier signature spanning immune and inflammatory signalling, tissue homeostasis and neurodevelopmental processes, extending beyond established markers of neurodegeneration.

In contrast, male *APOE* ε4 carriers showed enrichment for innate immune processes, MAPK signalling, receptor tyrosine kinase pathways, developmental biology, and growth factor signalling. Several male-specific predictors, including MMP12, TGFBI, SCARF2, KLK4, C1QL2 and KHK, showed preservation across cohorts. KLK4, which ranked among the strongest predictors in UKB and was the highest-ranked predictor in GNPC, has previously been implicated in plasma proteomic signatures of Alzheimer’s disease, dementia and cognitive decline (37). Although KLK4 is highly expressed in prostate tissue, whether this contributes to its prominence in the male *APOE* ε4 carrier signature remains unclear. MMP12 and TGFBI further highlighted extracellular matrix remodelling and tissue repair, with MMP12 previously associated with future dementia risk (38,39). C1QL2 (40) and KHK (41) pointed towards synaptic organisation and metabolic regulation, respectively. Overall, the male carrier signature emphasised tissue remodelling, cellular communication and metabolic processes.

Although immune-related pathways were represented in both profiles, their underlying components differed, with broader immune and inflammatory signalling featuring in female carriers and innate immune and MAPK-related signalling in male carriers. Consistent with the importance of sex-specific molecular investigations, a recent study of steroid hormones in Alzheimer’s disease identified distinct sex-stratified associations, including elevated cerebrospinal fluid cortisol in females with AD and elevated plasma aldosterone in males with AD (42).

Several proteins captured biological processes shared across female and male *APOE* ε4 carriers in UKB, with varying degrees of preservation across cohorts. BCAN and CDCP1 were identified among the top predictors in both female and male *APOE ε4* carriers in UKB and remained highly ranked when the UKB-trained models were applied to GNPC, supporting the preservation of these shared molecular signals across cohorts. In conventional association analyses, BCAN was the only predictor shared between female and male *APOE* ε4 carriers that showed significant associations with dementia outcomes (all-cause dementia and AD), with these associations observed exclusively among female carriers. BCAN encodes brevican, a brain-specific extracellular matrix proteoglycan involved in synaptic stabilisation, neuronal plasticity and maintenance of perineuronal nets, and disruption of extracellular matrix organisation has been increasingly implicated in Alzheimer’s disease and cognitive decline (43). CDCP1 has been linked to systemic inflammation, immune activation, and blood-brain barrier dysfunction (44), processes increasingly recognised as important contributors to dementia pathogenesis. Although its role in Alzheimer’s disease remains incompletely understood, the consistent identification of CDCP1 across *APOE* ε4 carrier groups suggests that inflammatory mechanisms may contribute to *APOE* ε4-associated molecular vulnerability. APOE was also identified among the top predictors in both carrier groups in UKB, although its cross-cohort replication was retained only in female carriers. APOE plays a central role in lipid transport and cholesterol homeostasis and represents one of the strongest genetic risk factors for Alzheimer’s disease (26). These findings suggest that *APOE* ε4-associated dementia risk may involve altered lipid metabolism alongside disruptions in extracellular matrix organisation and cellular communication.

Despite substantial heterogeneity across strata, GFAP, NEFL and GDF15 were consistently identified among the most important predictors across all four groups in UKB. This finding is consistent with a previous large-scale UK Biobank proteomic analysis (19), which identified these proteins among the strongest predictors of incident dementia. GFAP and NEFL are well-established biomarkers of astroglial activation and neuronal injury respectively and have repeatedly demonstrated strong associations with incident dementia, Alzheimer’s disease, and cognitive decline (45,46). GDF15 has emerged as a marker of biological ageing, cellular stress, mitochondrial dysfunction, and systemic inflammation, processes increasingly recognised as contributors to neurodegeneration (19). The consistent identification of these proteins across strata suggests that neurodegeneration, glial activation, and age-related biological stress represent common pathways underlying dementia risk irrespective of sex and genetic background.

Although NEFL and GDF15 remained common across both *APOE* ε4 carrier sex strata in GNPC, GFAP showed less consistent preservation. This may partly reflect differences between the UKB and GNPC cohorts, particularly the contrast between incident dementia prediction in initially dementia-free individuals and prevalent Alzheimer’s disease classification in a clinically enriched cohort. Differences in statistical power may also contribute to this finding. Plasma GFAP reflects reactive astrogliosis and has been shown to change early in the AD continuum, with elevations observed in cognitively unimpaired individuals with amyloid pathology and years before clinical symptom onset (45–47). Its relationship with AD pathology may also vary across disease stages. In contrast, NEFL reflects neuronal injury, while GDF15 captures broader processes related to ageing and cellular stress, which may remain informative across more heterogeneous disease contexts. NEFL and GFAP were significantly associated with all dementia-related outcomes across all sex and genotype strata. GDF15 was significantly associated with all three outcomes in female carriers and non-carriers, with VD in male carriers, and with all-cause dementia and AD in male non-carriers (**Supplementary Tables 40,42).**

In UKB, SYT1 was among the top predictors in both female and male *APOE* ε4 carriers, as well as female non-carriers, and conventional association analyses demonstrated significant associations with dementia outcomes in most strata. *SYT1* encodes synaptotagmin-1, a presynaptic vesicle protein involved in calcium-dependent neurotransmitter release and increasingly recognised as a marker of synaptic dysfunction in Alzheimer’s disease. Previous studies (48) have reported altered cerebrospinal fluid synaptotagmin-1 concentrations in both preclinical and symptomatic Alzheimer’s disease, supporting its role as a marker of early synaptic pathology. Therefore, SYT1 may reflect a broader synaptic component of dementia risk rather than an *APOE* ε4- or sex-specific signal.

A major strength of this study is the cross-cohort and cross-platform preservation of molecular feature importance, with SHAP correlations of approximately 0.70 in both sexes despite differences in cohort composition, disease stage, outcome definition (incident all-cause dementia versus prevalent AD) and proteomic platform (Olink versus SomaScan).

Although the highest-ranking predictors across nearly all subgroups were proteins, direct comparison between modalities should be interpreted cautiously given differences in feature numbers and correlations among molecular predictors. The presence of lipid-related measures among male non-carriers may indicate differential contributions of metabolic pathways across biological risk strata. Future studies incorporating additional lipidomic and metabolomic platforms may clarify these relationships.

Classifiers excluding age retained broadly comparable discrimination to models including age, although small improvements in AUC were observed with age inclusion in some subgroups. Given that age is the strongest established risk factor for dementia (2), the preservation of discrimination in models excluding age suggests that the identified omics signatures retain substantial predictive information without chronological age as an explicit predictor.

Current biomarker frameworks are largely applied uniformly across populations despite growing evidence that sex and *APOE* ε4 status modify Alzheimer’s disease biology and progression. The identification of partially distinct molecular signatures years before incident dementia across sex–*APOE* ε4 strata suggests that early risk prediction, biomarker interpretation, and potentially future therapeutic targeting may benefit from more biologically stratified and personalised approaches. The particularly robust molecular signature observed in female *APOE* ε4 carriers, alongside distinct replicated biological profiles in male carriers, further illustrates how stratification may reveal molecular signals that could be obscured when populations are analysed collectively.

This study has several limitations. First, the analyses were observational. Therefore, the identified molecular signatures should be interpreted as predictors and associations rather than evidence of mechanistic causality. Second, pathway enrichment analyses were exploratory, with no pathways surviving false discovery rate correction; therefore, pathway-level interpretations remain hypothesis-generating. Nevertheless, the consistency of broader biological themes across SHAP interpretation, enrichment analyses and epidemiological association testing supports the plausibility of the identified sex-specific molecular patterns. Finally, KNN imputation was performed before cross-validation, potentially introducing information leakage and optimistic bias in performance estimates. However, a targeted sensitivity analysis in a subset of female *APOE* ε4 carriers using revised preprocessing showed comparable discrimination and highly correlated predictions, suggesting limited sensitivity to these changes in the evaluated subset.

This study demonstrates that dementia risk is characterised by partially distinct molecular signatures across sex–*APOE* ε4 strata, with *APOE* ε4 carriers exhibiting stronger and more reproducible molecular patterns than non-carriers. The preservation of molecular feature-importance patterns when UKB-trained models were applied to an independent cohort, despite differences in disease stage and proteomic platform, supports their potential biological relevance and transportability. These findings underscore the importance of stratified approaches for future biomarker discovery, risk prediction and precision medicine strategies in neurodegenerative disease.

## Methods

### Population (UK Biobank)

The main cohort of the study was the UK Biobank (UKB). This is a large-scale prospective study including over 500,000 participants in the UK, aged 40–69 years at baseline. For this study, we were interested in investigating participants with both proteomic and metabolomic data. UKB Proteomic data was generated using the Olink® Explore 3072 assay (49), and metabolomic data was generated using the Nightingale Health® NMR platform. To identify the overlap between proteomic and metabolomic data, the two datasets were linked at the individual level using the unique UK Biobank participant identifier (eid). Before quality control, 53,029 participants had proteomics and 488,082 had metabolomics. Only participants with both proteomic and metabolomic measurements at baseline were retained. After merging the two datasets, the population of interest was 52,505 participants. Prevalent dementia cases were removed (n = 55).

Molecules with 25% or more missing data were excluded (n = 4): CST1, GLIPR1, NPM1, and PCOLCE. In addition, participants with 25% or more missing values across the retained study variables were excluded. After excluding prevalent dementia cases and applying the missingness criteria, 44,236 participants remained, comprising 1,167 incident dementia cases and 43,069 controls. Those lost to follow-up were removed as well as participants with no *APOE* genotype information, resulting in a final sample of 42,605 controls and 1,154 dementia cases with a median follow-up of 15 years (**Extended Data Figure 1**).

For stratification analyses, the dataset was split by sex and *APOE* ε4 status, resulting in 23,704 females (*APOE* ε4 carriers n = 7,007) and 20,055 males (*APOE* ε4 carriers n = 5,750). **Supplementary Table 2** includes the details of the sample numbers.

### Outcomes

The outcome of interest was all-cause dementia, which included all subtypes of dementia recorded in the UKB. Dementia cases were identified from linked electronic health records (hospital inpatient records, primary care records, and death certificate registers). Diagnosis dates were defined as the earliest qualifying dementia diagnosis from these data sources. The median follow-up duration was 14.6 years (IQR: 13.8–15.3 years).

### Predictors

The key predictors consisted of 3,244 omics (2,917 proteomics and 327 metabolomics). In addition, age, number of *APOE* ε4 alleles, ethnicity, years of education and BMI (body mass index) were included based on previous machine learning studies in dementia research (15,16,18,50,51). For the UKB Olink data, normalised protein expression (NPX) values provided by UKB were used directly, as these values are pre-normalised and log2-transformed by the assay pipeline. The complete list of proteins used can be found in **Supplementary Table 3.**

Metabolomic profiling was performed using high-throughput nuclear magnetic resonance (NMR) spectroscopy by Nightingale Health Ltd. The platform quantified a broad range of circulating metabolites spanning multiple biological pathways, including lipids and lipoproteins, amino acids, glycolysis-related metabolites, ketone bodies, and other low-molecular-weight compounds (52).

To minimise technical variation, metabolite measurements were processed using the standard technical correction procedure implemented in the ukbnmr R package. This approach applies robust linear regression-based correction to the 107 directly measured metabolites to account for known technical confounders, after which derived biomarkers and metabolite ratios are recalculated from the adjusted values. Additional metabolite ratios proposed within the ukbnmr framework and considered to have potential biological relevance were also included in the present analyses.

A total of 327 metabolite variables were included in the study, comprising 107 directly measured metabolites, 61 composite biomarkers, 81 predefined metabolite ratios, 76 additional metabolite ratios derived using the ukbnmr framework, and two additional Nightingale biomarkers (spectrometer-corrected alanine and glucose-lactate). The complete list of metabolite measurements is provided in **Supplementary Table 4.**

To facilitate reproducibility, UK Biobank field identifiers for variables included in the analyses are provided in **Supplementary Table 1a**.

### Pre-processing pipeline

Given the high dimensionality of the dataset and computational constraints, imputation was performed once on the full dataset prior to model development. Missing values were imputed using k-nearest neighbours (k=5), implemented with the KNNImputer class from scikit-learn (53). The choice of *k* was guided by prior studies applying similar methods to high-dimensional biomedical data (18,54). All classifiers were trained and evaluated on the imputed dataset.

Continuous variables were standardised to a mean of zero and a standard deviation of one within the nested cross-validation, using the *StandardScaler* class from scikit-learn. Standardisation was retained to harmonise feature scales for downstream analyses.

### Stratification

To investigate subgroup-specific molecular risk signatures, the UKB population was stratified by sex and *APOE* ε4 carrier status. Sex was defined using UK Biobank Field 31 (female or male), which was obtained from the NHS central registry at recruitment and, in some cases, subsequently updated by participants. *APOE* ε4 carriers were defined as individuals with at least one *APOE* ε4 allele (ε2/ε4, ε3/ε4 or ε4/ε4). This resulted in four strata: female *APOE* ε4 carriers, female non-carriers, male *APOE* ε4 carriers and male non-carriers. The machine learning pipeline was fitted independently for each stratum.

## Statistical analysis

### Cohort descriptive statistics

Baseline characteristics were summarised for the overall cohort and stratified by dementia diagnosis (cases vs. controls). Continuous variables were summarised as median and interquartile range (IQR), while categorical variables (sex, ethnic group, BMI category, and number of *APOE* ε4 alleles) were summarised as counts and percentages. Differences between participants with and without dementia were assessed using the Mann–Whitney U test for continuous variables and Pearson’s chi-squared test for categorical variables. Two-sided p-values were reported for descriptive purposes. Statistical analyses and table generation were performed using Python (SciPy, pandas, and python-docx libraries).

### Inferential statistics for *APOE* ε4 carrier status, sex, and dementia diagnosis

A multivariable logistic regression model was fitted with dementia diagnosis as the binary outcome (0 = no dementia, 1 = dementia) using the statsmodels package (v0.14.4) in Python v3.12.9. The main predictors were *APOE* ε4 carrier status, sex (binary coded), and their interaction term (*APOE* ε4 carrier × sex). Sex was coded as female = 0 and male = 1, while *APOE* ε4 carrier status was coded as non-carrier = 0 and carrier = 1, with female non-carriers serving as the reference group. Models were adjusted for age at baseline, years of education, BMI category, and ethnicity (categorised as White, Black and Other/Unknown, with the latter comprising South Asian, Other, Mixed, Chinese and Unknown/NA). This analysis was intended to assess the interaction between sex and *APOE* ε4 carrier status on the odds of incident dementia during follow-up, rather than to estimate time to dementia onset.

Variance inflation factors (VIFs) were calculated to assess multicollinearity among predictors. Odds ratios (ORs) and 95% confidence intervals (CIs) were obtained by exponentiating model coefficients. Additional contrast-based ORs were computed using linear hypothesis testing (model.t_test) to estimate subgroup-specific effects (e.g., *APOE* ε4 carriers vs. non-carriers stratified by sex).

### Machine learning pipeline

XGBoost is an ensemble tree-based machine learning framework built on gradient boosting principles, where sequential decision trees iteratively minimise prediction error by correcting residual errors from preceding trees (55). The method is particularly well suited to high-dimensional biomedical data because it can efficiently model nonlinear relationships and complex interactions between variables while remaining robust to complex feature structures (55). In addition, XGBoost is compatible with SHAP (SHapley Additive exPlanations) based interpretation, enabling biologically interpretable feature importance analyses.

Given these properties and its strong performance in prior dementia studies (56–59), XGBoost was selected as the classifier in this study.

Independent XGBoost pipelines were developed separately for each subgroup: Female *APOE* ε4 carriers, Female non-carriers, Male *APOE* ε4 carriers, and Male non-carriers. For each subgroup, two independent modelling pipelines were developed using the same preprocessing, feature selection, hyperparameter optimisation and evaluation procedures, differing only in whether age was included as a candidate predictor. This resulted in eight subgroup-specific pipelines. This comparison was performed to assess the extent to which molecular signatures retained predictive discrimination without chronological age as an explicit model predictor.

The data were split into a training dataset and a testing dataset. The split, stratified by dementia incidence, was 70/30 i.e., 70% of the data was used for training and 30% to test the classifiers. The methodology applied to each pipeline is described in detail below.

### Feature selection

To reduce dimensionality from over 3,000 features, we applied the Boruta algorithm (BorutaPy implementation) (60) with a Random Forest classifier as the base estimator, separately for each sex-*APOE* ε4 subgroup. Categorical variables were converted to categorical type and one-hot encoded; boolean variables were converted to binary format (0/1).

Boruta was applied to the training data only to avoid data leakage. Training was performed within a repeated stratified 5-fold cross-validation framework (with 3 repeats), using n_estimators=’auto’ to allow BorutaPy to determine the number of trees automatically, perc=100 to retain only features with importance greater than the most important shadow feature (minimising false positives), and a maximum of 50 iterations to balance stability with computational efficiency. Balanced class weights were used to address class imbalance.

A feature was deemed stable if selected in ≥40% of all cross-validation runs. Stable features for each subgroup were retained for subsequent machine learning model development, with the selected feature set held fixed throughout nested cross-validation.

### Classifier development

For each sex–*APOE* ε4 subgroup, we trained gradient-boosted tree models using XGBoost within a repeated nested cross-validation framework (outer 5-fold CV; inner 3-fold CV for hyperparameter tuning; repeated 5 times). Models were fit on the Boruta-selected feature set (stability ≥ 0.40), retrieved from the original dataset and preprocessed within the final modelling pipeline (continuous variables standardised using a StandardScaler fitted on the corresponding model-fitting data, categorical variables one-hot encoded, and boolean variables converted to binary). Class imbalance was addressed using scale_pos_weight, computed as the ratio of negative to positive cases in the training dataset. All models used GPU acceleration with histogram-based tree growth (tree_method=’hist’, device=’cuda’).

Hyperparameters were optimised by randomised search (10 iterations, ROC-AUC scoring) over predefined ranges (**Supplementary Table 5**). For each subgroup, the best hyperparameters selected by the inner 3-fold cross-validation within each outer training fold were aggregated across the 25 outer-fold iterations (5 folds × 5 repetitions) to obtain a single configuration for final model fitting, reducing dependence on any individual training partition. Numerical hyperparameters (e.g., learning rate, subsample) were aggregated by taking the mean, while categorical hyperparameters (e.g., booster type) were aggregated using the mode. During nested cross-validation, decision thresholds were selected separately using predictions from each outer validation fold to achieve specificity ≥ 0.85 while maximising recall. The mean threshold across outer folds was used to generate the aggregated cross-validation confusion matrix.

The final XGBoost model was retrained on the training set using the aggregated hyperparameters derived from nested cross-validation. For probability calibration, the training data were further split into sub-training (80%) and calibration (20%) subsets, stratified by dementia status; the XGBoost classifier was refitted on the sub-training subset and subsequently calibrated using sigmoid regression (CalibratedClassifierCV, method = “sigmoid”, cv = “prefit”), fitted on the calibration subset. For the final calibrated model, the decision threshold was selected on the calibration subset using the same criterion (specificity ≥ 0.85 while maximising recall) and subsequently applied to the held-out test set. Model performance was evaluated on the independent held-out test set using calibrated probabilities. Performance metrics included discrimination (ROC-AUC), calibration (Brier score), accuracy, specificity, recall, precision, and F1 score. Threshold-dependent metrics and confusion matrices were calculated using the previously selected calibration-derived threshold.

### Sensitivity analysis

To assess potential data leakage arising from KNN imputation performed before the training– test split, a targeted sensitivity analysis was conducted in a subset of 2,000 female *APOE* ε4 carriers. The analysis used 68 previously selected features from models excluding age as a predictor and a fixed XGBoost configuration. Both preprocessing approaches were evaluated using the same stratified 70/30 training–test split. The original approach, in which KNN imputation was fitted to the full dataset before splitting, was compared with a revised approach in which imputation was fitted exclusively on the training data and subsequently applied to the test data. Model discrimination was evaluated using AUC, and agreement between predicted probabilities was assessed using Pearson’s correlation coefficient.

### Bootstrap evaluation

Two complementary bootstrap procedures were applied. First, for each subgroup’s final model, uncertainty in performance metrics (ROC-AUC, recall, precision, F1 score, specificity, and accuracy) was quantified by stratified bootstrap resampling of the held-out test set with replacement (1,000 replicates). Dementia cases and controls were resampled separately, retaining the original sample size of each group in every replicate. Point estimates were paired with 95% percentile confidence intervals derived from the bootstrap distributions.

Second, to directly compare discrimination between the with-age and no-age pipelines, paired nonparametric bootstrap resampling was performed on the same held-out test set (1,000 stratified replicates). Within each replicate, cases and controls were resampled with replacement, and the same bootstrap indices were applied to both models before recomputing ROC-AUC. The difference in AUC (ΔAUC; AUCwith-age − AUCno-age) was recorded across replicates. We report the observed ΔAUC and its 95% percentile bootstrap confidence interval, calculated from the 2.5th and 97.5th percentiles of the bootstrap distribution.

Bootstrap resampling was selected because it does not rely on parametric distributional assumptions and enables robust estimation of uncertainty for performance metrics, as well as paired comparison of models evaluated on the same individuals.

### Shap Feature interpretation

Feature importance and interpretation were computed using SHAP (SHapley Additive exPlanations) (61), implemented with shap.Explainer applied to the trained XGBoost estimator. SHAP values were calculated on the transformed held-out test set, using the correspondingly transformed training set as the background dataset. Global feature importance was summarised as the mean absolute SHAP value for each feature, and features were ranked accordingly. SHAP summary and bar plots were generated to visualise the direction, magnitude, and relative importance of feature contributions. For reproducibility, we report the SHAP details used in **Supplementary Table 6**.

### SHAP-Based Gene Set Enrichment Analysis

Pathway enrichment analyses were conducted as an exploratory approach to identify biological themes associated with sex-specific differences in UKB SHAP-derived protein importance rankings. Enrichment was performed using a pre-ranked Gene Set Enrichment Analysis (GSEA) approach implemented in the clusterProfiler and ReactomePA packages in R (v4.4.0). To investigate biological processes associated with sex differences in model-derived protein importance, separate female-versus-male contrasts were generated for *APOE* ε4 carriers and *APOE* ε4 non-carriers.

For each comparison, proteins were ranked using mean absolute SHAP (SHapley Additive exPlanations) values obtained from the subgroup-specific UKB XGBoost models. Protein importance values from the female and male models were merged using the union of proteins retained in either model. Proteins absent from a subgroup-specific model were assigned a SHAP importance of zero, reflecting their absence from that fitted model. This approach was intended to compare the complete model-derived protein signatures, capturing differences in both feature selection and relative SHAP importance between sexes, rather than estimating intrinsic sex-specific biological effects. To account for differences in the distributions of SHAP importance between independently trained models, mean absolute SHAP values were standardised within each sex using z-score transformation. This allowed the subsequent contrast to capture differences in relative feature importance between the sex-specific predictive signatures, rather than differences in absolute SHAP magnitudes. A contrast score was then calculated for each protein as the difference between the standardised female and male SHAP values (female minus male), such that positive values indicated greater relative importance in females and negative values indicated greater relative importance in males.

Protein symbols were mapped to Entrez Gene identifiers using the org.Hs.eg.db annotation database. Where multiple proteins mapped to the same Entrez identifier, the protein with the largest absolute contrast score was retained. Proteins were then ranked according to their contrast scores to generate a pre-ranked gene list for enrichment analysis.

Gene Set Enrichment Analysis was performed using the gseGO() function against Gene Ontology Biological Process (GO:BP) terms and the gsePathway() function against the Reactome pathway database, using nPermSimple = 10,000. Multiple testing correction was performed using the Benjamini–Hochberg procedure. Only gene sets containing between 5 and 500 genes were considered.

Enrichment results were summarised using normalised enrichment scores (NES), where positive NES values indicate pathways enriched among proteins with greater relative importance in females and negative NES values indicate pathways enriched among proteins with greater relative importance in males. Results were visualised using custom dot plots displaying pathway enrichment direction, gene set size, and false discovery rate-adjusted significance.

### External replication of the proteomic signal (GNPC)

To evaluate cross-platform transportability of the UKB proteomic signal, external validation was performed using the Global Neurodegeneration Proteomics Consortium (GNPC) v1 harmonized dataset (21), accessed through the Alzheimer’s Disease Data Initiative (ADDI) AD Workbench (62). GNPC is a harmonised multi-cohort proteomics resource enriched for neurodegenerative disorders, including AD, Parkinson’s disease (PD), amyotrophic lateral sclerosis (ALS), and frontotemporal dementia (FTD). External validation analyses were restricted to *APOE* ε4 carrier subgroups, as these models showed the strongest performance in UKB. In addition, the models chosen for validation were the models not including age as a predictor given the robust performance in UKB. Only GNPC baseline visits were included in the present analysis.

This external validation framework represents a stringent test of cross-cohort transportability due to substantial differences between datasets, including disease stage (incident dementia in UKB versus prevalent dementia in GNPC), outcome definition (all-cause dementia in UKB versus clinically diagnosed AD in GNPC), proteomic platform (Olink in UKB versus SomaScan in GNPC), and cohort age structure.

### GNPC proteomic platform

Proteomic measurements in GNPC were generated using SomaScan SOMAmer technology (versions 3, 4, and 4.1), comprising approximately 7,000 aptamers per sample, whereas UKB proteomics were measured using the Olink platform. Preprocessing and harmonisation of the GNPC v1 dataset were performed as previously described by the consortium (21). In this study, we standardised GNPC protein abundance values using z-score normalisation, centring each protein to mean zero and scaling to unit variance. Cohort-specific standardisation was applied to harmonise feature scales while avoiding transfer of distributional parameters across proteomic platforms.

### GNPC proteomic predictors and feature harmonisation

Neither carrier-specific model retained metabolomic predictors following feature selection; therefore, no metabolite imputation was required for external validation in GNPC. UKB Olink protein predictors were mapped to GNPC SomaScan proteins by protein symbol. Where multiple SomaScan aptamers corresponded to a single protein, aptamer values were collapsed using the median abundance across aptamers. For UKB model predictors not measured in GNPC, a constant value corresponding to the UKB training-set mean was initially assigned to all GNPC participants, thereby preserving the original model input structure. Following GNPC-specific z-score standardisation, these constant-valued predictors became undefined (NaN) due to zero variance and were handled by XGBoost’s native missing-value mechanism. This pragmatic approach enabled application of the original UKB models without refitting. Feature ordering was matched exactly to the original UKB training matrices separately for female and male *APOE* ε4 carrier models before prediction.

### GNPC outcome variable

The primary outcome in GNPC was AD diagnosis. Cases were defined as individuals with AD diagnosis (AD = 1). Controls were defined as individuals without AD diagnosis (AD = 0) and with a Clinical Dementia Rating (CDR) score <0.5. Individuals not meeting either definition were excluded from analysis.

### GNPC classifier

Final UKB-trained XGBoost classifiers were exported in UBJSON format and applied directly to GNPC without retraining. The female *APOE* ε4 carrier model retained 1,000 boosting rounds and 68 input features, while the male *APOE* ε4 carrier model retained 1000 boosting rounds and 71 input features, confirming preservation of the original UKB-trained architectures. Predicted probabilities were generated after harmonising GNPC predictors to the corresponding UKB feature order. Classification thresholds previously optimised and calibrated in UKB were transferred unchanged to GNPC for external evaluation. Performance metrics included ROC-AUC, recall, specificity, precision, F1 score, and accuracy.

### SHAP analysis and feature concordance

To assess biological concordance across cohorts, SHAP values were computed for the externally validated GNPC predictions using the UKB-trained XGBoost models. Global feature importance was summarised using mean absolute SHAP values. Concordance between UKB and GNPC feature importance rankings was assessed using Spearman correlation across shared proteins. Overlap among the top-ranked proteins between cohorts was additionally quantified, and concordance plots were generated to visualise preservation of feature importance structure across datasets.

### *APOE* ε4 carrier replicated predictors associations with all-cause dementia, AD and vascular dementia (UKB only)

Exploratory, post-hoc association analyses were conducted to further characterise the relationships between proteins identified as replicated predictors across UKB and GNPC and multiple dementia-related outcomes within UKB APOE ε4 carrier populations, stratified by sex. As protein selection was partly informed by predictive importance within the same cohort, these analyses were intended to provide complementary epidemiological characterisation rather than independent confirmatory evidence.

Specifically, proteins consistently ranked among the top 20 predictors in both UKB and GNPC for each sex-specific *APOE* ε4 carrier group were selected for these association analyses. GFAP was additionally included in both sex-specific analyses because it was consistently identified across all UKB sex and *APOE* ε4 strata and has been widely implicated in early neurodegeneration (45,46). APOE and SYT1 were also included in the male carrier analysis because they ranked among the top predictors in the corresponding UKB model and replicated in female carriers.

Analyses were performed separately in female and male *APOE* ε4 carriers using multivariable logistic regression implemented in Python (v3.12.9) with pandas (v2.2.3), numpy (v2.1.3), and statsmodels (v0.14). For each sex-specific analysis, only individuals carrying one or two *APOE* ε4 alleles were included. Three binary outcomes were evaluated independently: all-cause dementia, Alzheimer’s disease (AD), and vascular dementia (VD). These outcomes were not mutually exclusive, and participants with multiple dementia diagnoses could contribute as cases to more than one analysis. To ensure outcome specificity, a shared “healthy control” group was defined as individuals free from all three diagnoses simultaneously (Dementia = 0, AD = 0, and VD = 0). For each outcome, cases were compared against this common control population, to avoid contamination of controls with alternative dementia diagnoses.

Proteomic predictors were analysed individually, with a separate multivariable logistic regression model fitted for each protein-outcome combination. Protein abundance values were standardised to z-scores prior to analysis, while continuous covariates (age at baseline and years of education) were similarly z-score normalised. *APOE* ε4 allele count (1 or 2 copies) was modelled as a continuous gene-dose covariate, and categorical covariates (ethnic_group and BMI_levels) were encoded using categorical dummy variables. Models were adjusted for age at baseline, *APOE* ε4 allele count (included as a numeric gene-dose covariate), years of education, ethnicity, and BMI category.

Robust HC3 standard errors were used to reduce sensitivity to heteroscedasticity. Associations were summarised as odds ratios (ORs) with 95% confidence intervals (CIs). Multiple testing correction was performed separately within each sex-specific analysis and dementia outcome, across the proteins tested, using the Benjamini–Hochberg false discovery rate (FDR) procedure. Statistical significance was defined as FDR q < 0.05. Results were visualised using heatmaps of log₂(OR), volcano plots, and forest plots of FDR-significant associations.

### *APOE* ε4 non-carrier secondary association analyses (UKB only)

To further investigate the association of shared predictors across all strata with multiple dementia outcomes, sex-stratified multivariable logistic regression analyses were additionally conducted in the *APOE* ε4 non-carrier populations following the same analytical framework described in the previous Section. GFAP, GDF15 and NEFL were selected because they were consistently identified among the top predictors across all UKB sex and *APOE* ε4 strata. SYT1 protein was additionally included because it was identified among the top predictors in all UKB strata except male non-carriers. These analyses aimed to determine whether the shared predictors identified through the prediction models were also associated with multiple dementia outcomes in conventional association analyses for the non-carrier population.

## Code availability

Code and scripts for the analysis can be found in the following GitHub repository: https://github.com/saracalhas/sex_apoe4_dementia_prediction

## Data Availability

The UK Biobank data used in this study are available to eligible researchers through the UK Biobank application process, subject to approval and the UK Biobank data access procedures. This research was conducted using the UK Biobank Resource under application number 78867. The GNPC v1 Harmonized Dataset used for external validation is available to qualified researchers through the Alzheimer's Disease Data Initiative AD Workbench, subject to the GNPC Data Use Agreement and Publication Policies.

## Acknowledgements

We are grateful to the participants of the UK Biobank. The National Health Service (NHS) Northwest Centre for Research Ethics Committee approved the UK Biobank study (reference 11/NW/0382), which conforms to the provisions of the Declaration of Helsinki. We thank the participants, contributing cohorts, investigators and staff of the Global Neurodegeneration Proteomics Consortium (GNPC), whose clinical and proteomic data made the external validation in this study possible. The harmonized GNPC V1 dataset was accessed and analysed through the Alzheimer’s Disease Data Initiative AD Workbench, and data discovery and analysis services were provided in-kind by the AD Data Initiative. We acknowledge Gates Ventures and Johnson & Johnson for supporting the majority of GNPC biosample proteomic analyses, as described in the GNPC resource publication.

We also thank the High-Performance Computing cluster (Apocrita) at QMUL for facilitating these analyses, and all members of the Centre for Preventive Neurology for their ongoing support. The Centre for Preventive Neurology is supported by a grant from Barts Charity. Sara Calhas is supported by the Wellcome Trust PhD programme – Health Data in Practice: Human-Centred Science (218584/Z/19/Z). Petroula Proitsi is supported by Alzheimer’s Research UK grant ARUK-PPG2024-012.

