## Extended Data Figures for "Sex shapes plasma molecular signatures of dementia risk across *APOE* ε4 strata"

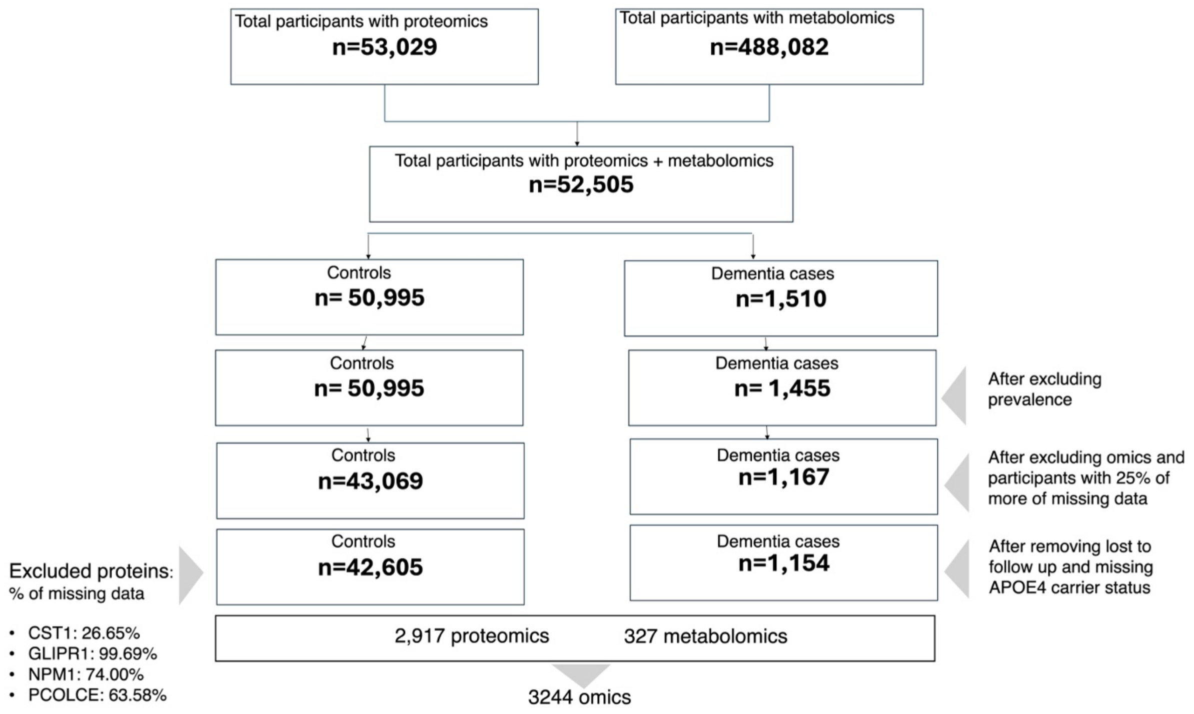


**Extended Data Figure 1**. Cohort selection flow diagram. UK Biobank participants with baseline proteomics (Olink® Explore 3072) and metabolomics (Nightingale Health® NMR) data. Exclusion criteria included ≥25% missingness across omics features, ≥25% missingness at the participant level, prevalent dementia at baseline, loss to follow-up, and unavailable genotype information. The final cohort comprised 42,605 controls and 1,154 incident dementia cases, with a median follow-up of 15 years.

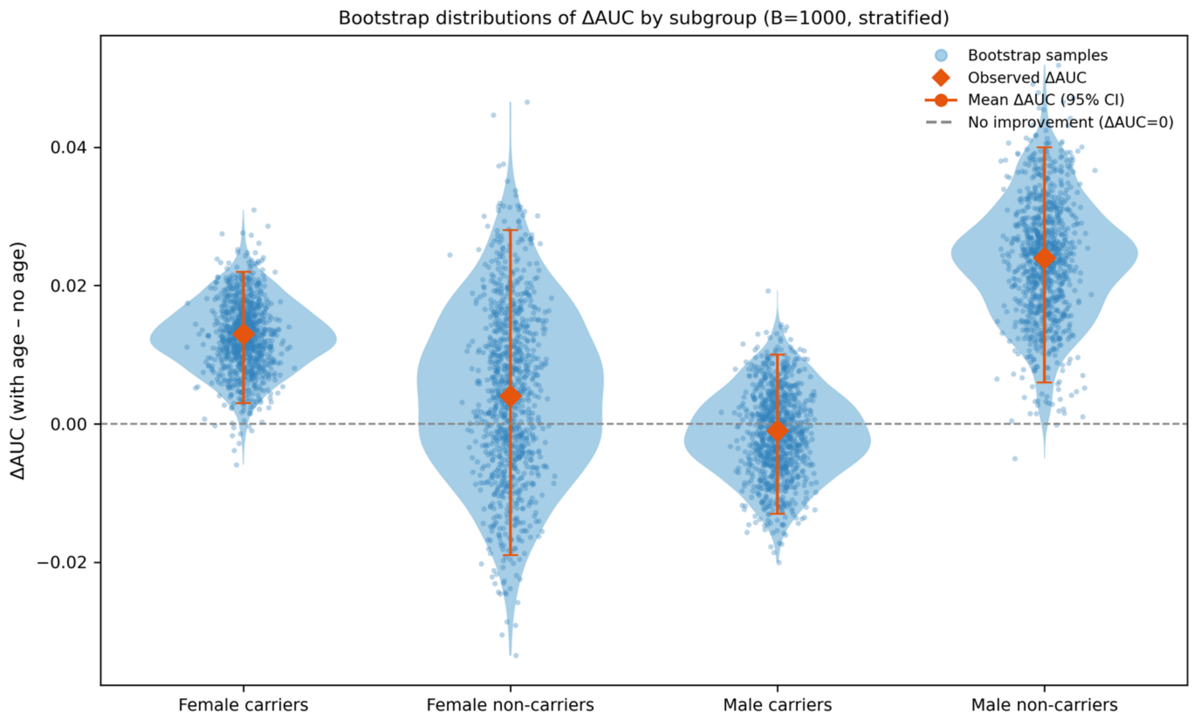


**Extended Data Figure 2.** Violin plot showing bootstrap distribution of AUC difference of classifiers per APOE4 carrier status with and without age included as predictor alongside the molecules.

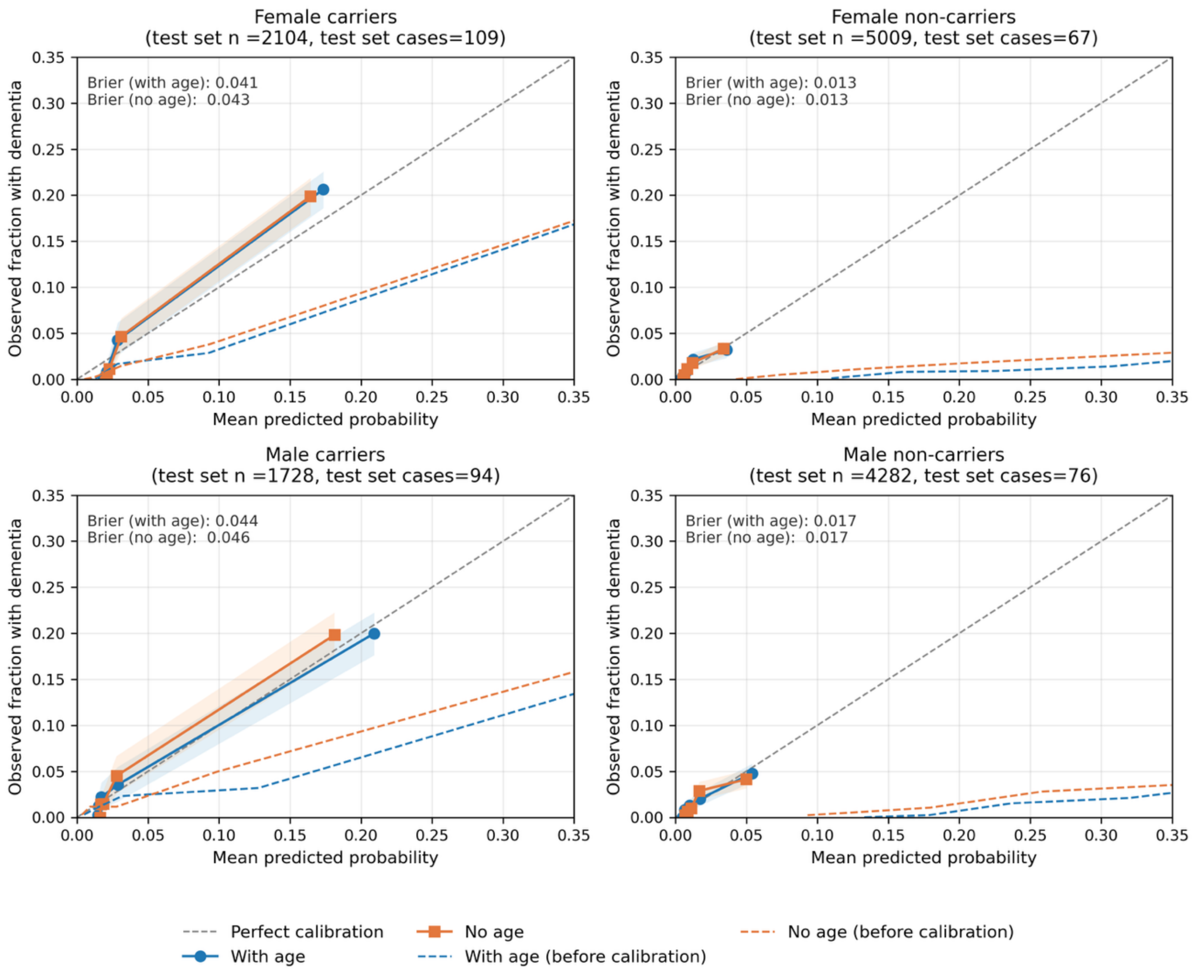


**Extended Data Figure 3.** Calibration plots for dementia classifiers stratified by sex and APOE4 status, with and without age included as a predictor. The solid lines show observed versus predicted probabilities after sigmoid calibration, with dashed grey lines indicating perfect calibration. Dotted lines show uncalibrated predictions for reference. Numbers indicate Brier scores (lower = better). Sigmoid calibration improved agreement between predicted and observed risks across subgroups, with little difference between models with and without age.

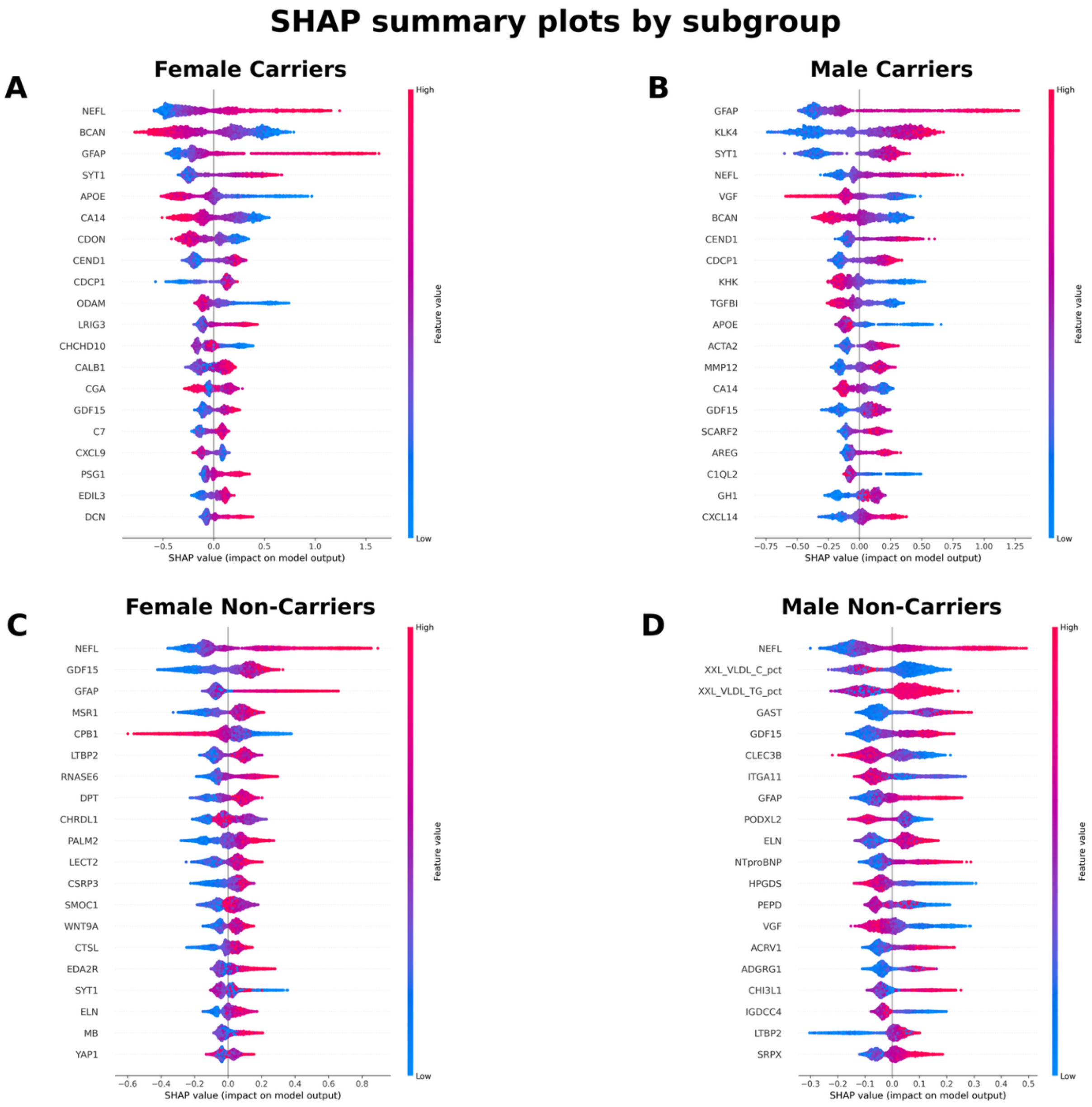


**Extended Data Figure 4. SHAP summary plots for dementia classifiers stratified by sex and APOE4 carrier status (no-age models).** Panels show the top features ranked by mean absolute SHAP value for (A) female carriers, (B) male carriers, (C) female non-carriers, and (D) male non-carriers. Points represent individual participants, with horizontal position indicating SHAP value (feature contribution to predicted dementia risk) and colour representing feature value (blue = low, red = high).

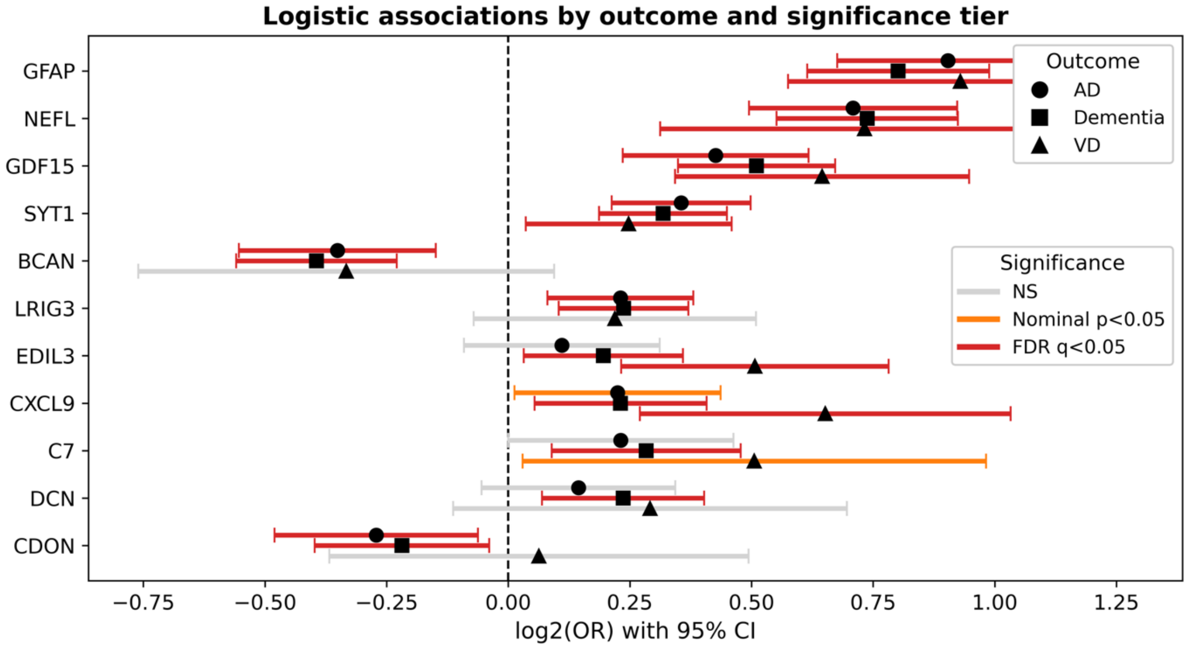


**Extended Data Figure 5**. Sex-stratified logistic regression associations of replicated female APOE ε4 carrier predictors with all-cause dementia, Alzheimer's disease (AD), and vascular dementia (VD) in UK Biobank. Points represent $\log_2$ odds ratios and error bars indicate 95% confidence intervals from multivariable logistic regression models adjusted for age, education, APOE ε4 allele count, BMI category, and ethnicity. Colour indicates significance level after multiple testing correction. Only proteins reaching significance (n=11) are shown.

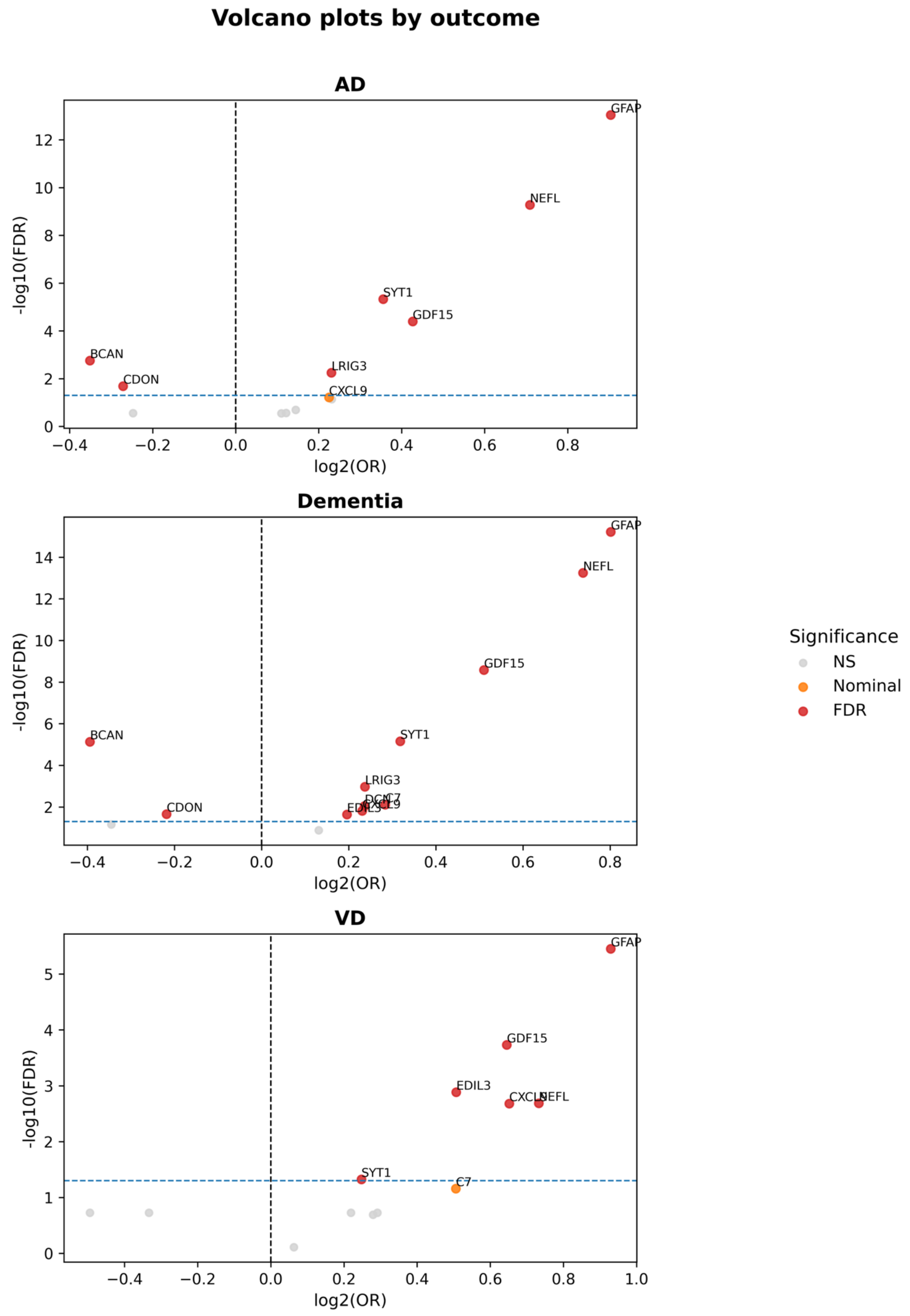


**Extended Data Figure 6**. Volcano plots showing sex-stratified logistic regression associations of replicated female APOE ε4 carrier predictors with Alzheimer's disease (AD), all-cause dementia, and vascular dementia (VD) in UK Biobank. The x-axis represents $log_2$ odds ratios and the y-axis shows $-log_{10}$(FDR-adjusted p-values). Points are coloured according to significance level after multiple testing correction. GFAP and NEFL demonstrated the strongest positive associations across outcomes, while BCAN and CDON showed inverse associations primarily with AD and all-cause dementia.

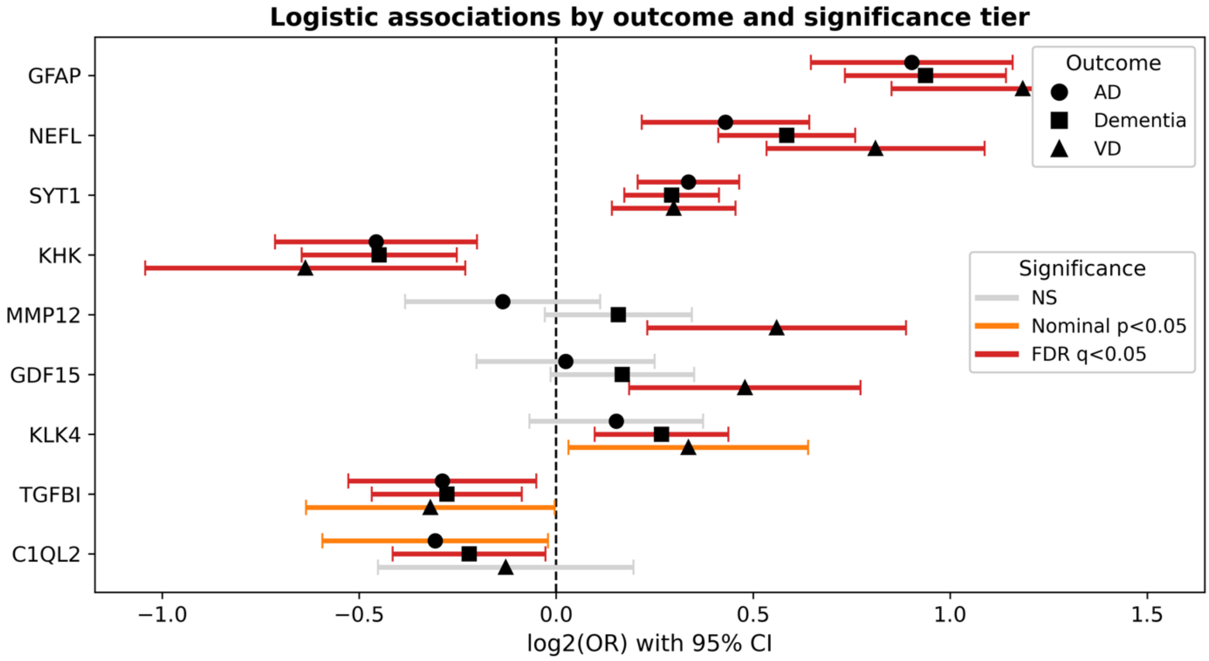


**Extended Data Figure 7**. Sex-stratified logistic regression associations of replicated male APOE ε4 carrier predictors with all-cause dementia, Alzheimer's disease (AD), and vascular dementia (VD) in UK Biobank. Points represent $log_2$ odds ratios and error bars indicate 95% confidence intervals from multivariable logistic regression models adjusted for age, education, APOE ε4 allele count, BMI category, and ethnicity. Colour indicates significance level after multiple testing correction. Only proteins reaching significance (n=9) are shown.

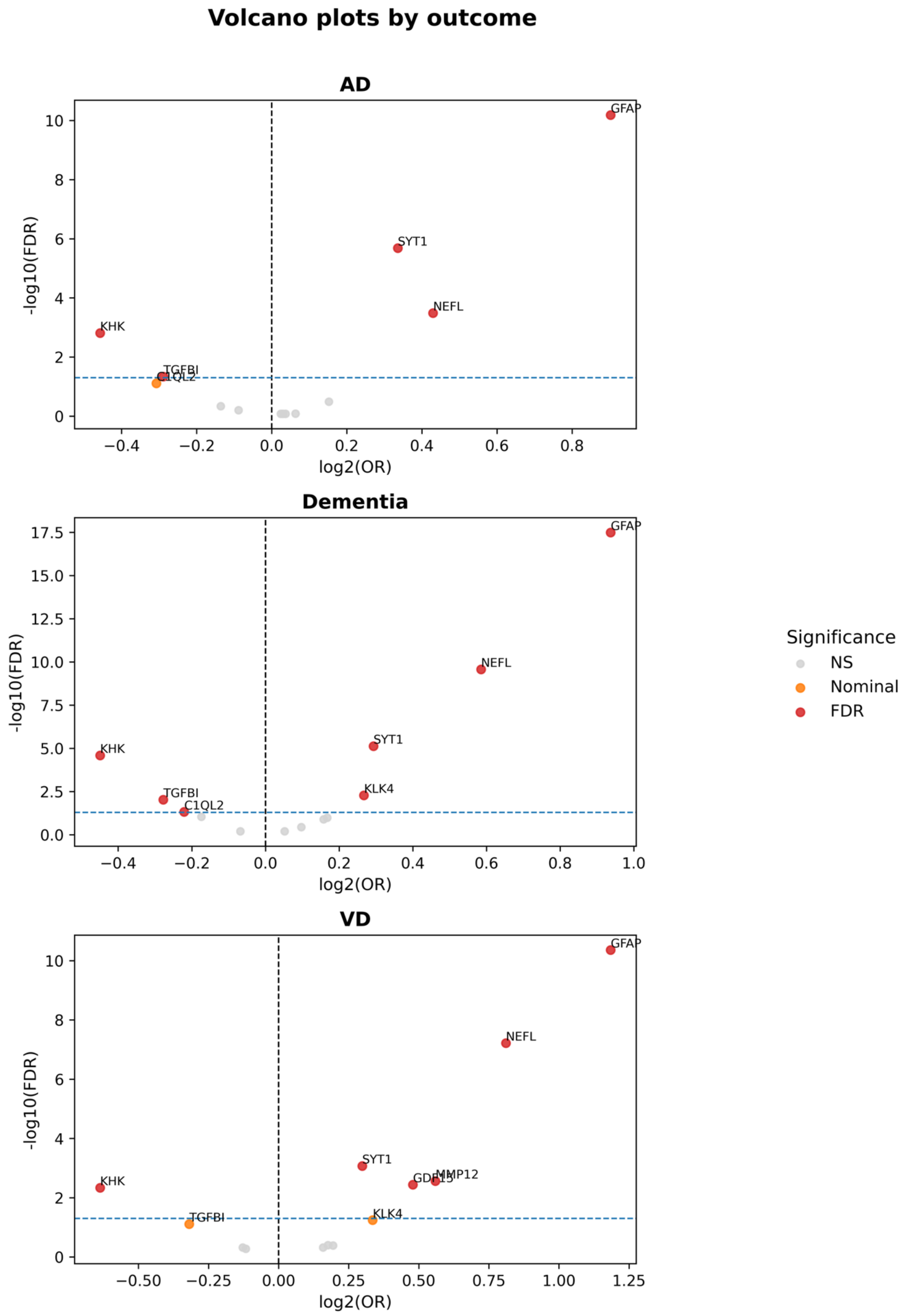


**Extended Data Figure 8.** Volcano plots showing sex-stratified logistic regression associations of replicated male APOE ε4 carrier predictors with Alzheimer's disease (AD), all-cause dementia, and vascular dementia (VD) in UK Biobank. The x-axis represents $\log_2$ odds ratios and the y-axis shows $-\log_{10}$(FDR-adjusted p-values). Points are coloured according to significance level after multiple testing correction. GFAP and NEFL demonstrated the strongest and most consistent positive associations across dementia outcomes, while GDF15, C1QL2 and MMP12 showed outcome-specific associations.
